# Super-resolution imaging pinpoints ultrastructural changes at the node of Ranvier in patients with polyneuropathy

**DOI:** 10.1101/2022.08.05.22278366

**Authors:** Luise Appeltshauser, Janis Linke, Hannah S. Heil, Christine Karus, Joachim Schenk, Katherina Hemmen, Claudia Sommer, Kathrin Doppler, Katrin G. Heinze

## Abstract

To shed light on nanoscale pathologies in patients with polyneuropathy, we assessed human nerve biopsies by super-resolution fluorescence microscopy. We focused on both physiological protein arrangement and pathological ultrastructural changes at the node of Ranvier, a crucial region of the peripheral myelinated axon. Direct stochastic optical reconstruction microscopy (*d*STORM) revealed a ∼190 nm periodic protein arrangement of cytoskeletal proteins and axoglial cell adhesion molecules. Periodic distances increased at the paranodal region of the node of Ranvier in patients with polyneuropathy, both at the axonal cytoskeleton and at the axoglial junction. In-depth image analysis of human nerve biopsies revealed a partial loss of proteins of the axoglial complex (Caspr-1, neurofascin-155) in combination with detachment from the cytoskeletal anchor protein ß2-spectrin. Super-resolution dual-color colocalization data was supported by high-content confocal imaging combined with deep learning-based analysis, indicating that paranodal elongation occurs especially in acute and severe axonal neuropathy, as a possible correlate of Wallerian degeneration and related cytoskeletal damage. Our findings show that super-resolution imaging can identify, quantify and map elongated periodic protein distances in peripheral nerve biopsies for pathophysiological studies and direct implications for diagnostic assessment.

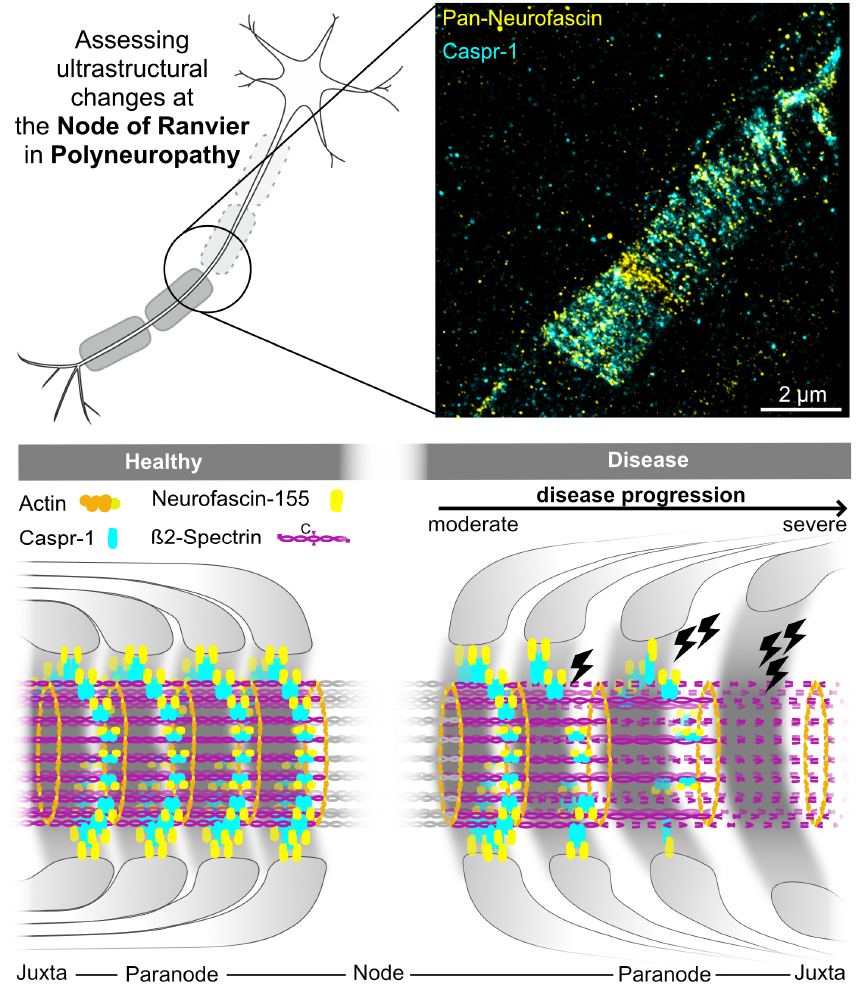

**Graphical abstract:** **Pathological alterations of ultrastructural protein arrangement at the paranodal region of the node of Ranvier in polyneuropathy**.Super-resolution microscopy allows for assessing ultrastructural protein arrangement and pathological alterations in patients with polyneuropathy. In human healthy nodes, axoglial and axoskeletal proteins follow a 190 nm periodic arrangement (left). In pathologically altered nodes (right), periodic protein distances of axonal ß2-spectrin elongate, in combination with elongation and partial loss of the axoglial complex of Caspr-1 and neurofascin-155. The axoglial complex itself colocalizes closely even in pathologically altered nodes. A complete loss of the axoglial complex could be the ultrastructural correlate of the detachment of paranodal myelin loops. Scale bar 2 μm.

## Introduction

Polyneuropathies are the most common disorder of the peripheral nervous system (1). They are characterized by impaired signal transduction of peripheral nerves, resulting in severe sensorimotor deficits with high disability in affected patients (2). Different pathomechanisms involve axonal, myelin and Schwann cell damage. In recent years, the essential role of the node of Ranvier and the severe consequences of its alterations have come to focus (3, 4).

The node of Ranvier of myelinated axons is the key element for rapid long-distance neuronal signal transduction via saltatory conduction (5–7). Its highly organized nanostructure allows for clustering ion channels at the nodal region and securing cell adhesion at the paranodal and juxtaparanodal region (8–11). The complex Schwann cell - axon interplay is mediated by selected cell adhesion proteins at the paranode, like glial neurofascin-155 and axonal contactin-1 associated protein 1 (Caspr-1). They form part of the axoglial complex and attach the terminal myelin loops to the axon (6, 12, 13). In rodents, new super-resolution imaging techniques have shown that these cell adhesion molecules, and also ion channels and axonal scaffold proteins, are arranged following a ∼190 nm periodicity (14). Yet, little is known about the nanostructural protein arrangement of the node of Ranvier in humans, and especially in pathologically altered nerves.

Resolving such small protein periodicities would not be possible without the application of super-resolution microscopy, which provides an unpreceded insight into the molecular organization and protein interaction in health and disease (15). Both Stimulated Emission Depletion (STED) and direct Stochastic Optical Reconstruction Microscopy (*d*STORM) (16, 17) combine target specificity with a very high spatial resolution and allow analysis of periodic ultrastructures by computational autocorrelation analysis (18, 19). In contrast to STED, *d*STORM in particular pinpoints molecular positions in cells and tissue with nanometer precision. With thousands of hits per image, *d*STORM provides maps of the subcellular architecture as a prerequisite for further analysis of molecular clusters, colocalization and relative positions (20– 22). So far, super-resolution microscopy has mainly been applied to map the neuronal nanoscale anatomy under physiological (14, 23–26), but rarely under pathological conditions (27–29). Despite its potential for pathophysiological understanding and diagnostic assessment, *d*STORM has not been applied on human nerves.

Here, we apply *d*STORM to ex vivo human sensory nerve teased fiber preparations of patients with polyneuropathy, and analyze the nanoscale periodicity of selected adhesion and scaffold proteins at the node of Ranvier. To reveal underlying mechanisms of pathological alterations, we also assessed the interaction of the axoglial adhesion molecules neurofascin-155 and Caspr-1 and the axonal scaffold protein ß2-spectrin by dual-color colocalization analysis. To assess node of Ranvier morphometrics in parallel, a tailored high-content confocal imaging pipeline as a complementary modality was implemented. Considering histopathological acuteness, type and severity of the disease as well as clinical features in the analysis, we could relate our nanoscale findings to histopathological and clinical data, thereby exploring both pathomechanistic implications and the diagnostic potential of our methods.

## Results

### Definition and validation of histopathological categories of polyneuropathy

In order to assess the nodo-paranodal nanoscale organization in the human species and to identify possible pathological changes, we included n = 17 patients with polyneuropathy into our study. These had undergone sensory nerve biopsy (n = 16 sural nerve, n = 1 radial superficial nerve) during the routine diagnostic workup process. Demographic data and electrophysiological and clinical diagnosis including diagnostic criteria are listed in Supporting Table S1. An experienced examiner made the histopathological diagnosis from standard histological stains, and classified according to the primarily affected structure (axonal / demyelinating), the acuteness (acute / chronic) and the severity of axonal loss (mild / moderate / severe; see Supporting Table S2). Histopathological and clinical parameters differed significantly in acute vs. chronic and axonal vs. demyelinating neuropathy. Furthermore, electroneurographic features of the respective sensory nerve correlated to the histopathological severity of axonal loss. The nerve fiber density was significantly reduced in acute vs. chronic neuropathy (see Supporting Table S3 and Supporting Fig. S1). Accordingly, we separated our patients into three histopathological subcohorts: acute axonal neuropathy (n = 6 patients), chronic axonal neuropathy (n = 6 patients) and chronic demyelinating neuropathy (n = 5 patients). This allowed for a direct comparison of ultrastructural changes in nodo-paranodal nanoscale organization in relation to type, acuteness and severity of neuropathy.

### Development and validation of a *d*STORM autocorrelation analysis tool

Patient derived tissue preparations pose a whole set of challenges for super-resolution imaging. Especially a limited immunolabeling efficiency and nonspecific binding can compromise the image reconstruction quality and resolution. To retrieve the periodic arrangement of structural proteins in the internodal and paranodal region in patient derived tissue samples with *d*STORM microscopy, we developed a custom and easy-to-use image analysis tool implemented in MatLab (Mathworks Inc.). This tool is based on image autocorrelation of selected regions of interest within the super-resolved reconstructions, and allows for directly retrieving the periodicity of a certain protein ring structure from peak-to-peak distances in its respective autocorrelation profile. The general image processing workflow is illustrated in Supporting Fig. S2. We validated the performance and robustness of our approach using simulated datasets. To define a ground truth model for our simulations, we assumed a previously described 190 nm periodic arrangement of paranodal proteins (14) (Figure 1 A) and tested whether the image autocorrelation approach can achieve high levels of accuracy and robustness even in the challenging conditions presented in this study. As a result, we could reliably detect the ground truth of a 190 nm periodic arrangement in a simulated datasets despite high background levels and strongly reduced marker densities (Figure 1 B,C, and Supporting Fig. S3). Changes in the periodicity could be distinguished clearly, respectively (Figure 1 D,E).

**Fig. 1.**
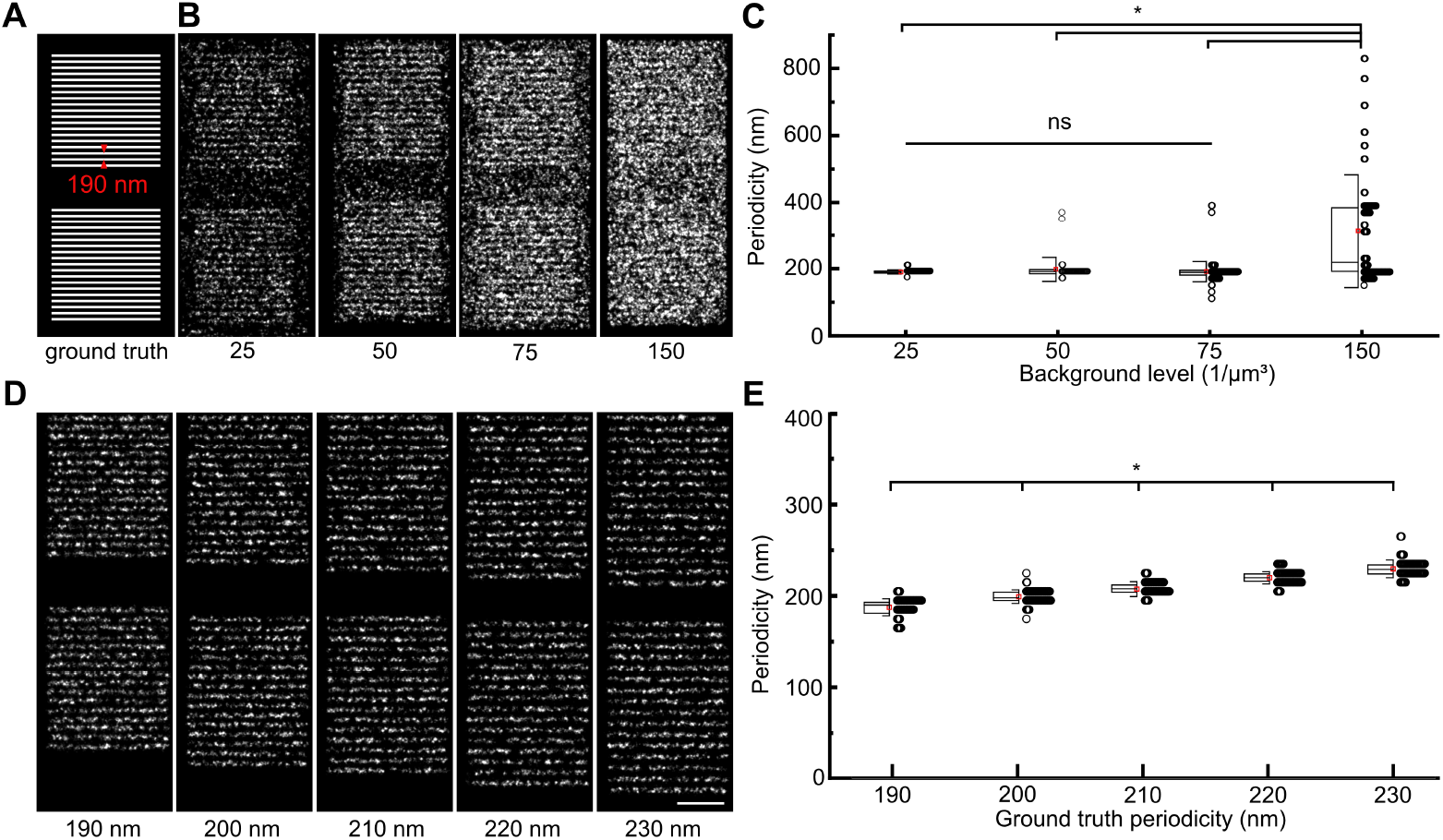
Super-resolution image autocorrelation analysis reliably identifies structural periodicity. (A) The robustness of image autocorrelation analysis is tested on simulated Single Molecule Localisation Microscopy (SMLM) data based on a line structure with 190 nm periodicity as a ground truth. (B) For background levels ranging from 25 - 150 events/μm^3^, autocorrelation recovers the underlying periodicity spacing as visualized in (C) boxplots, only failing at the highest background level (median ± standard deviation; 25: 190 ± 6 nm, n = 99; 50: 190 ± 36, n = 90; 75: 188 ± 30 nm, n = 143; 150: 218 ± 170 nm, n = 88; Kruskal-Wallis test with Dunn’s correction for multiple testing. (D) For ground truth structures with periodicities varying from 190-230 nm, autocorrelation (E) successfully retrieves and clearly distinguishes the different periodicity spacings (median ± standard deviation; 190: 187.6 ± 9.3 nm; 200: 199.1 ± 7.4 nm; 210: 207.5 ± 8.1 nm; 220: 219.9 ± 6.6 nm; 230: 229.6 ± 9.9 nm; n=60 in all cases, Kruskal-Wallis test with Dunn’s correction for multiple testing). Scale bar 1 μm.

### The adult human axonal internodal scaffold follows a ∼190 nm periodic arrangement

Next, we implemented the autocorrelation analysis on super-resolved *d*STORM image reconstructions of murine and human teased fiber preparations, significantly increasing the resolution in comparison to diffraction-limited microscopy (Figure 2A, inlet). First, we studied the periodicity of the internodal axoskeleton. In mice, we were able to display a periodic arrangement of ß2-spectrin, with a mean periodic distance of 188 nm, thus corresponding to previously described data (30). In human sural nerves, we found a mean periodicity of 184 nm, without significant difference to periodic distances in rodents (Figure 2 B and C, detailed values are shown in Supporting Table S4). Thus, the ∼190 nm structural protein periodicity is conserved in human adults.

**Fig. 2.**
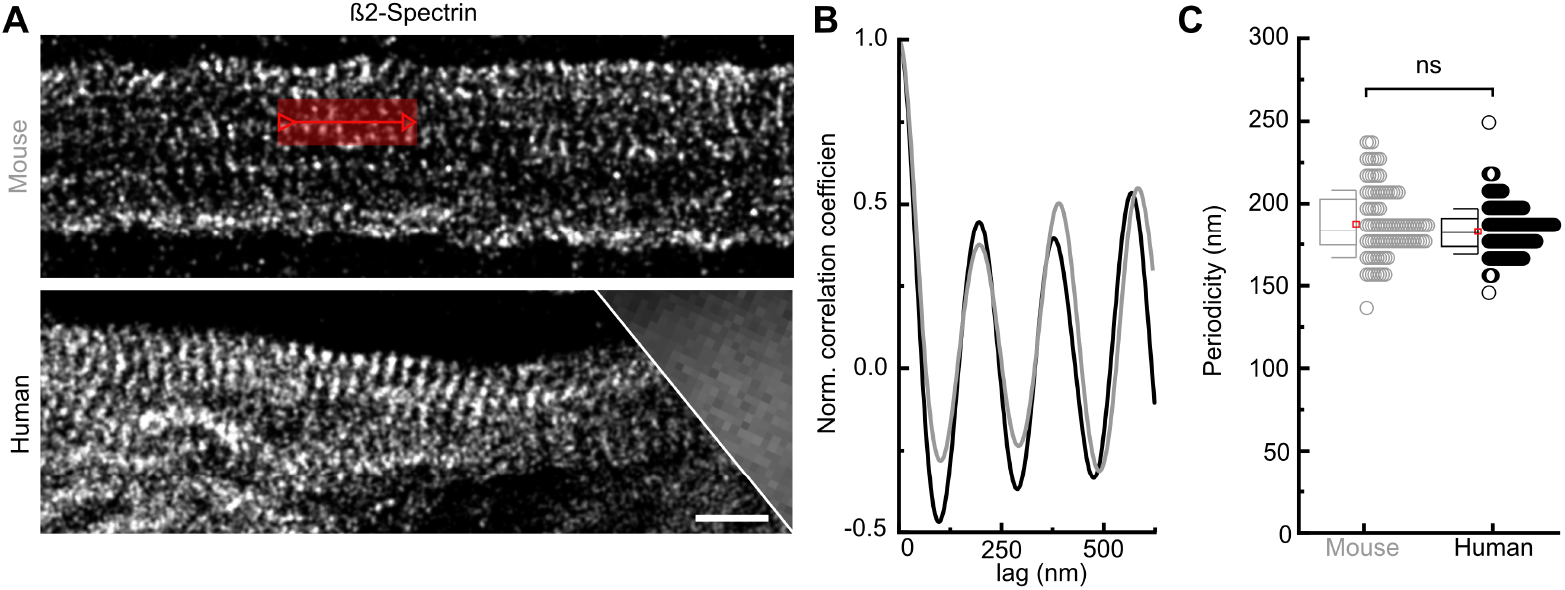
Super-resolution image autocorrelation analysis resolves the periodic cytoskeletal ultrastructure at the internode in rodents and humans. (A) *d*STORM resolves the longitudinal periodic arrangement (red arrow) of internodal ß2-spectrin in teased fiber preparations of murine (top) and human (bottom) axons, in contrast to conventional diffraction-limited widefield microscopy (inset, right). (B) Oscillating normalized correlation coefficient curves show equal ∼190 nm peak-to peak distances and thus an equal periodic arrangement of murine (gray) and human (black) axonal ß2-spectrin. (C) Boxplots of the distribution of periodic spacings measured by the autocorrelation analysis in murine (n = 99) and human teased fiber preparations (n = 192) show no significant differences in median periodic arrangement between the two species, tested by Mann-Whitney test. Boxplots display mean (red square), median (line), standard deviation (whiskers) and 25-75% (box). Scale bar 1 μm.

### Disruption of the ultrastructural periodic arrangements of paranodal proteins in patients with polyneuropathies

We then addressed the ultrastructural protein arrangement in the paranodal region of the node of Ranvier in rodents and humans. First, we analyzed the periodicity of axonal cytoskeletal ß2-spectrin in the paranodal compartment. In contrast to the internodal segment, the ultrastructural arrangement of ß2-spectrin was altered at the paranodal axon in patients with axonal neuropathies (see Figure 3 A, C, E). We could detect a significant increase of the periodicity of ß2-spectrin at the paranode in patients with axonal neuropathies compared to rodents (202 vs. 184 nm, p = 0.002). In patients with demyelinating neuropathies, however, the paranodal ß2-spectrin periodicity did not differ significantly from rodents (191 vs. 184 nm, p = 0.78). Differences in the periodicity between patients with axonal and demyelinating disease were not significant (see Supporting Table S4). Next, we assessed the paranodal axoglial junctional protein Caspr-1, located at the axon together with contactin-1 and tethered to axonal ß2-spectrin through protein 4.1B, and the glial binding partner of contactin-1, neurofascin-155 (9, 13, 31–33). The mean periodic distance of the axonal cell adhesion molecule Caspr-1 was severely elongated in patients with axonal and demyelinating neuropathy compared to rodents (247 ± 99 nm vs 187 ± 25 nm, p < 0.0001, see Figure 3 B,D,F and Supporting Table S4). The same applied for its glial counterpart neurofascin-155 (284 ± 125 nm vs. 173 ± 24 nm, p < 0.0001, see Figure 3 F and Supporting Table S4). In our patient samples, we observed a broad distribution of spacings that ranged from “healthy” nodes (with physiological periodic arrangement) to highly pathological nodes or heminodes (with disturbed periodicity). Figure 3 D illustrates that pathological alterations of the paranodal nanostructure follow a longitudinal direction, from the node to the internodal axon. This mid-to-lateral increase in periodic protein distance could be the correlate of a loosening of the helical Caspr-1 coils in pathologically altered nodes of Ranvier. In summary, *d*STORM was able to detect pathological alterations of the paranodal periodic protein arrangement in polyneuropathy in general, but especially on the level of individual nodes and patients, thus demonstrating its potential diagnostic aptitude to assess nanoscale pathology in selected patients.

**Fig. 3.**
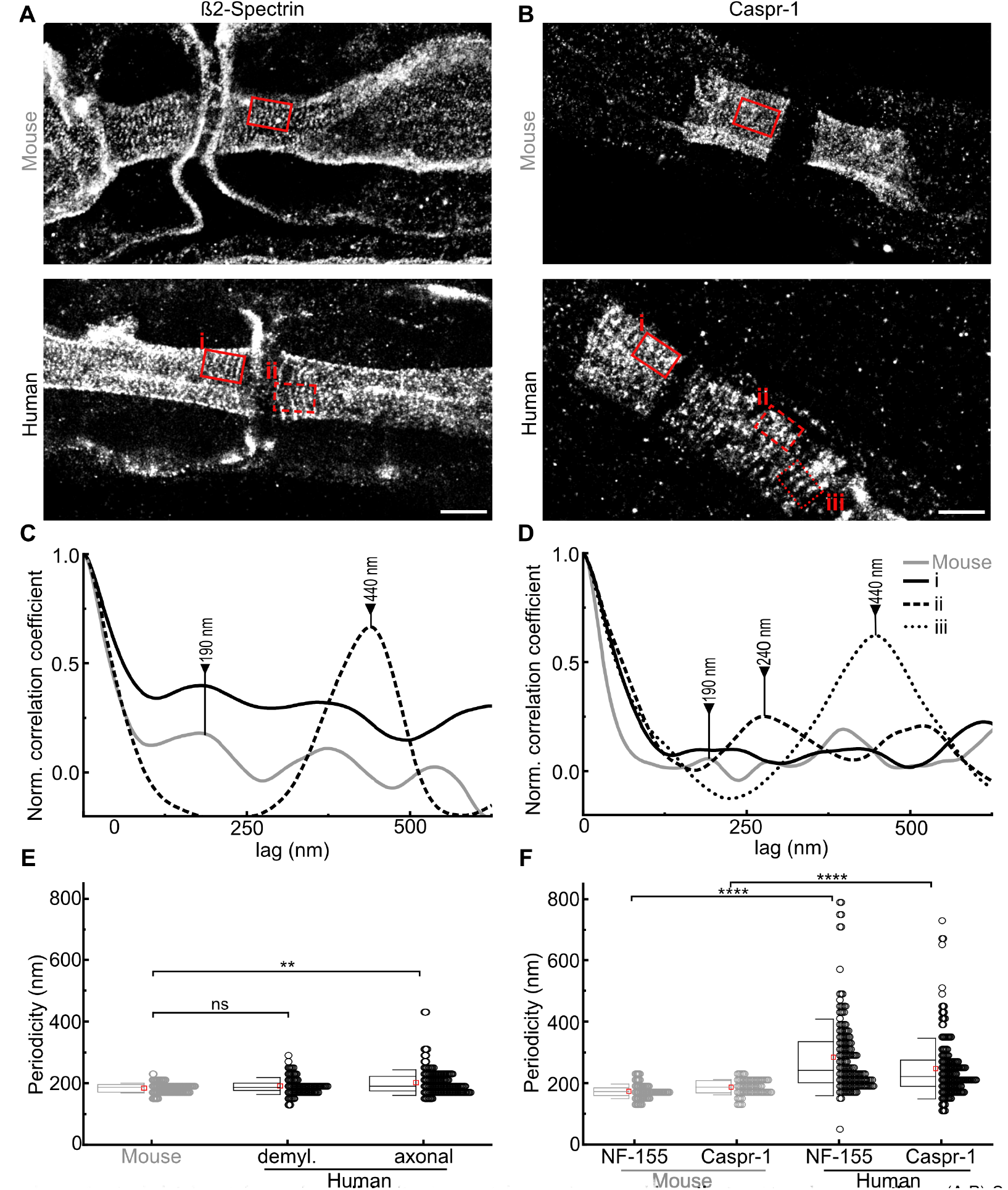
Periodic spacings of cytoskeletal proteins and axoglial adhesion proteins are increased in patients with polyneuropathies. (A-B) Super-resolved image reconstruction of (A) cytoskeletal ß2-spectrin and (B) axonal adhesional Caspr-1 organization at the node of Ranvier in teased fiber preparations of murine (top) and human (bottom) ex vivo axons. (C-D) Autocorrelation coefficient curves of the image regions marked in red in (A-B) of the distribution of ß2-spectrin (C) and Caspr-1 (D) in murine (gray lines) and human (black lines) teased fiber preparations indicating a local extension of the observed periodicity in pathological affected paranodes (dashed and dotted lines). (E) Boxplots of the distribution of periodic spacings of paranodal ß2-spectrin measured by autocorrelation analysis in mice (n = 130) and patients with histopathologically demyelinating (demyl., n = 106) and axonal (n = 159) neuropathy show a significant increase in cytoskeletal ß2-spectrin periodic arrangement in patients with axonal neuropathy, but not with demyelinating neuropathy. (F) Boxplots of the distribution of periodic spacings of Caspr-1 and neurofascin-155 (NF-155) in mice (n = 80 & 92) and in patients (n=194 & 173) show a highly significant increase in periodic spacings of axoglial proteins in patients with all types of polyneuropathies, with high variance between single regions of interest. Kruskal-Wallis test with Dunn’s correction for multiple testing was used for comparison. Significance level: * p < 0.05, ** p < 0.01, *** p < 0.001, **** p < 0.0001. Boxplots display mean (red square), median (line), standard deviation (whiskers) and 25-75% (box). Scale bars 2 μm.

### The axoglial complex remains attached even in pathologically altered nerves

We performed a colocalization analysis of Caspr-1, neurofascin-155 and ß2-spectrin on drift-corrected dual-color *d*STORM datasets of murine and human teased nerve fibers and quantitatively assessed colocalization with Manders’ A and B coefficients (34, 35). In mice, axonal Caspr-1 colocalized perfectly both with its glial counterpart neurofascin-155, as well as with its cytoskeletal anchor ß2-spectrin, represented by high Manders’ coefficients (Figure 4 A, B, E, F and Supporting Table S4). In human samples, on the other hand, colocalization of axonal Caspr-1 with the paranodal fraction of cytoskeletal ß2-spectrin was disrupted in patients with axonal and demyelinating polyneuropathies, represented by significantly reduced Manders’ A and B coefficients, with partial loss of Caspr-1 alongside alterations of the cytoskeletal periodic arrangement (Figure 4 D, F). Still, our data indicated an integrity of the paranodal axoglial complex of Caspr-1 and neurofascin-155, even if nodo-paranodal architecture was severely disrupted: Despite a partial loss of Caspr-1/ neurofascin-155 staining at the paranode, Manders’ A and B correlation coefficients did not decrease compared to murine samples in patients with demyelinating, chronic axonal and acute axonal neuropathies (see Figure 4 C, E and Supporting Table S4). In conclusion, our data indicate that axoskeletal alterations could contribute to lateral diffusion and loss of the axoglial complex at the paranodes, as the junctional complex itself remains closely attached even in pathologically altered nodes.

**Fig. 4.**
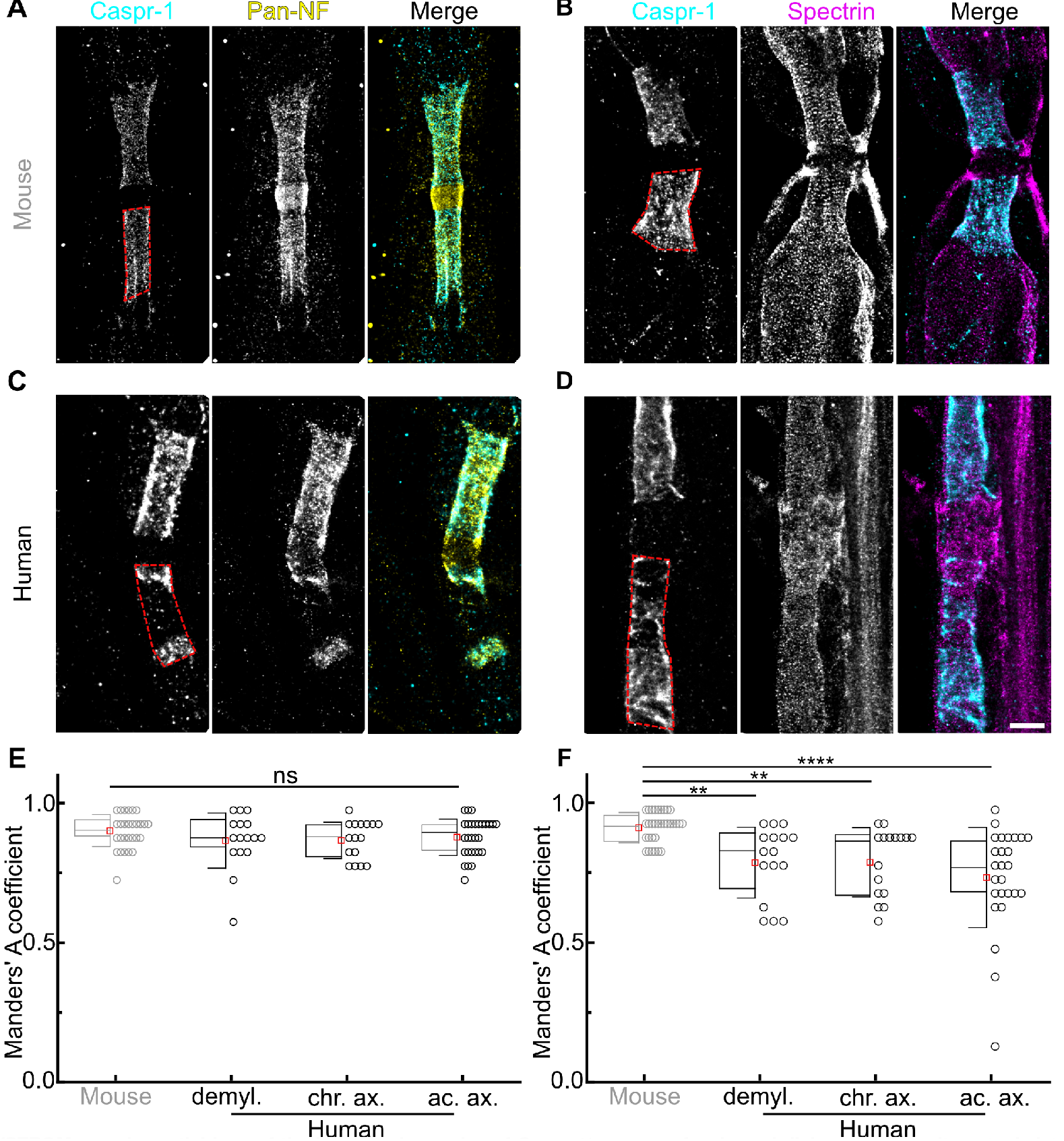
Dual-color *d*STROM reveals partial loss of the junctional complex of Caspr-1/pan-neurofascin and disjunction from its cytoskeletal anchor protein ß2-spectrin in polyneuropathy. (A-D) Super-resolved image reconstruction show colocalization of Caspr-1 (cyan) and pan-neurofascin (Pan-NF, yellow; A, C) and Caspr-1 and ß2-spectrin (magenta; B, D) in murine (A-B) and human (C-D) teased fiber preparations. Exact colocalization appears white. Gaps within the paranodal Caspr-1/panneurofascin staining illustrate partial loss of the axoglial complex at the paranode in a patient with acute and severe axonal neuropathy (C). Note: neurofascin-staining in the nodal region was not considered in the analysis, as the pan-neurofascin antibody stains both paranodal neurofascin-155 and nodal neurofascin-186 isoform. Analysis was performed within the paranodal region only (red dashed line). (D) In areas of loss of axonal Caspr-1, ß2-spectrin staining is still present, but pathologically altered without clearly visible periodic arrangement. (E) Boxplots show the distribution of Manders’ correlation coefficients between the axoglial complex Caspr-1 and neurofascin-155 in murine paranodes (n = 28) and paranodes of patients with demyelinating (demyl. n = 16), chronic axonal (chr.ax., n = 16) and acute axonal (ac.ax., n=28) neuropathy, without significant differences between the groups. (F) The distribution of Manders’ correlation coefficients between axonal junctional Caspr-1 and cytoskeletal anchor protein ß2-spectrin in the same groups (n = 32 / 16 / 16 / 26 paranodes) show significantly decreased colocalization and thus a detachment of Caspr-1 from its anchor molecule, especially in the group of acute axonal neuropathies. Testing was performed using Kruskal-Wallis with Dunn’s correction for multiple testing. Significance level: * p < 0.05, ** p < 0.01, *** p < 0.001, **** p < 0.0001. Boxplots display mean (red square), median (line), standard deviation (whiskers) and 25-75% (box). Scale bars 2 μm.

### High content confocal imaging reveals physiological nodo-paranodal parameters and identifies the node of Ranvier density as surrogate parameter for axonal loss

To relate our findings on a nanoscale level to the analysis of a higher number of nodes per patient, to “capture the bigger picture” of node of Ranvier pathology in patients with polyneuropathies of different etiologies and to evaluate the diagnostic potential of teased fiber immunofluorescence imaging, we performed high-content confocal imaging including large field of view photomicrographs on triple-stained teased fiber preparations and assessed the number of nodes of Ranvier and morphometrics in relation to type and stage of neuropathy. As data are scarce, we firstly defined normal values of nodo-paranodal parameters measured on sural nerves of human adults, as shown in Figure 5 A-B and the Supporting Movie M1. The mean nodo-paranodal length was 10.0 ± 4.9 μm, the mean nodal length in humans was 0.9 ± 0.3 μm and the axonal paranodal diameter was 2.3 ± 0.9 μm, without any significant differences to murine sural nerve parameters (see Table 1). These values can be considered as normal values of unaltered nodes of Ranvier in adult humans. Next, to check whether the teased fiber samples represented type and severity of neuropathy as assessed by cross-sectional gold standard analysis, we measured the node of Ranvier density per tissue volume on each sample by manual counting and by the application of deep learning strategies (see Supporting Notes 2 and 3). Counting accuracy using a fully automated deep learning approach was 1.21 ± 0.77 in human samples, with a highly significant correlation to manual assessment (see Supporting Note 3 and Figure S4). Patients with acute axonal neuropathy showed a significantly reduced nodal count compared to patients with chronic axonal neuropathy and chronic demyelinating neuropathy (16.1 vs. 33.3 vs. 35.9, p = 0.010 and p = 0.037, see Figure 5 C). We found a strong correlation of the total count of nodes of Ranvier in our teased fiber samples with parameters assessed by gold standard cross-sectional analysis: Teased fiber node density correlated with the semi-quantitative rating of histopathological severity of axonal loss (Spearman r = -0.75, p < 0.0001), the mean fiber density as an exact quantification of histopathological severity of axonal loss (Spearman r = 0.68, p = 0.002, see Figure 5 D) and with the mean count of Wallerian degeneration figures (Spearman r = -0.54, p = 0.026). Thus, we confirmed that our teased fiber preparations were representative of the histopathological diagnoses. Furthermore, we showed that the node of Ranvier density in teased fiber preparation is a possible surrogate marker for the severity of axonal loss that can be assessed accurately in human samples for diagnostic purposes by the application of deep learning strategies.

**Fig. 5.**
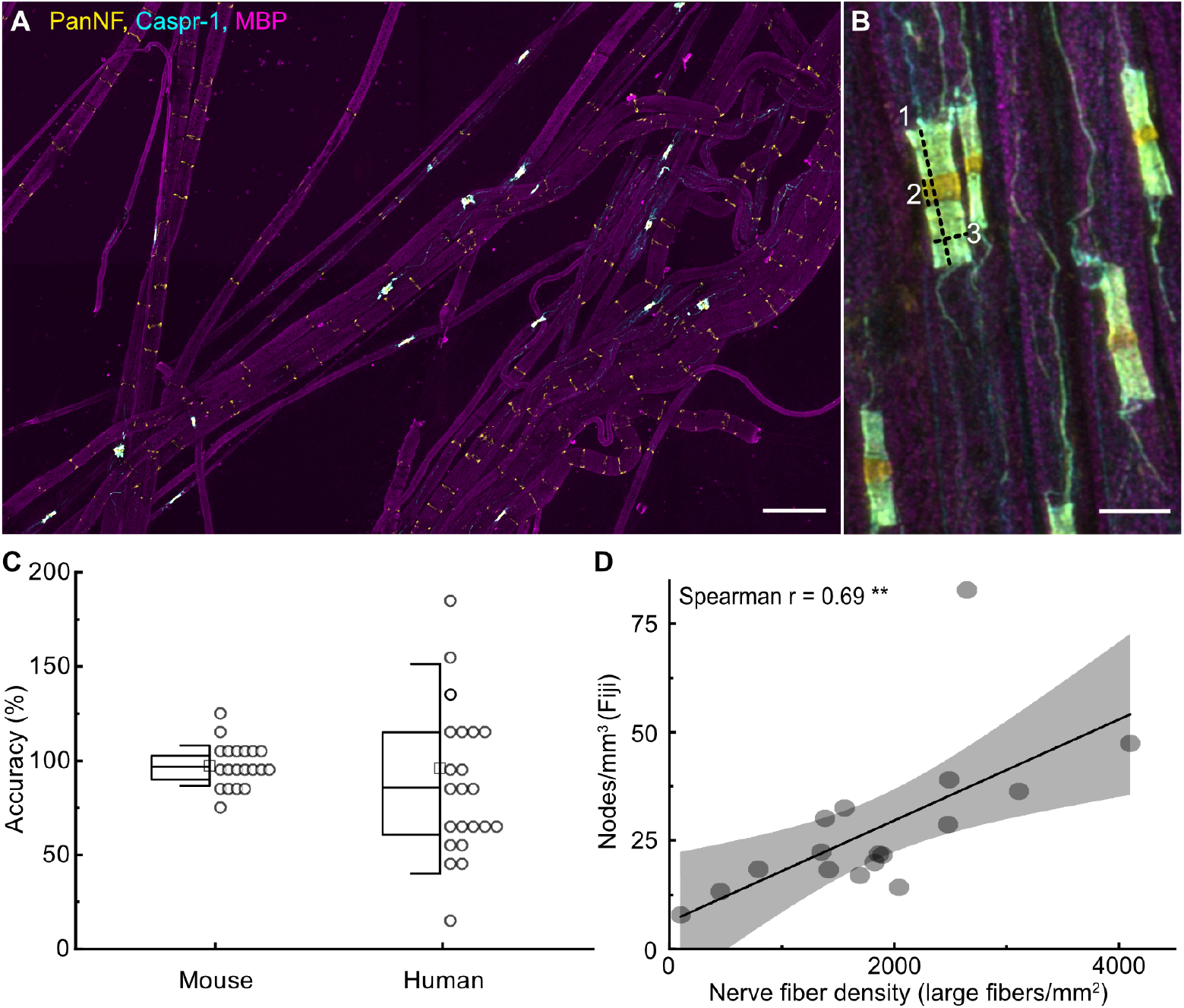
Assessing nodo-paranodal morphometrics and nodal count in large field of view photomicrographs. (A) large field of view photomicrographs of bundle teased fiber preparations of murine sural nerves show healthy myelinated fibers (MBP staining, magenta) with nodes of Ranvier expressing neurofascin-155 and neurofascin-186 (pan-neurofascin, yellow) and Caspr-1 (cyan), with colocalization at the paranode (white). Scale bar = 50 μm. (B) Nodo-paranodal morphometrics including nodo-paranodal length (1), nodal length (2) and axonal diameter (3) were assessed as indicated. Scale bar = 5 μm. (C) Accuracy of automated vs. manual node of Ranvier counting are shown for murine samples and human samples. In murine samples, automated analysis resulted in highly accurate results, whereas accuracy in human samples was reduced due to pathological node of Ranvier alterations. (D) The scatter plot shows a positive correlation of the teased fiber node of Ranvier density assessed manually with FIJI and the nerve fiber density assessed by gold-standard cross-sectional analysis in the whole cohort (Pearson r = 0.69, p = 0.002). Significance level: * p < 0.05. ** p < 0.01, *** p < 0.001, **** p < 0.0001.

**Table 1.** Nodo-paranodal morphometrics in murine and human sensory nerves - physiological and pathological conditions. The table displays means ± standard deviations (SD) of nodo-paranodal parameters (nodo-paranodal length, nodal length and paranodal axonal diameter) measured on Caspr-1 staining in human sural nerve intact nodes of Ranvier, and nodes of Ranvier of patients classified as acute axonal, chronic axonal and chronic demyelinating neuropathy (>180 nodes per cohort). Adjusted p-values are displayed compared to intact human nodes using Kruskal-Wallis test with Dunn’s correction for multiple testing. Individual patient values and n numbers are displayed in Supplemental Table S5. Significance levels: * p < 0.05, ** p < 0.01, *** p < 0.001, **** p < 0.0001. Abbreviations: n.a = not applicable, n.s. = not significant, SD = standard deviation.

| Category | node/tissue volume (1/mm<sup>3</sup>) | paranodal axonal diameter (μm) | nodo-paranodal length (μm) | nodal length (μm) |
| --- | --- | --- | --- | --- |
| intact human nodes | n.a. | 2.3 ± 0.9. | 10.0 ± 4.9. | 0.9 ± 0.3. |
| acute axonal neuropathy | 16.1 ± 3.5 | 2.3 ± 0.9<br>n.s. | 15.4 ± 10.1<br>*** | 1.3 ± 1.4<br>n.s. |
| chronic axonal neuropathy | 33.3 ± 10.0 | 2.2 ± 0.9<br>n.s. | 12.6 ± 7.1<br>**** | 1.2 ± 2.0<br>n.s. |
| chronic demyelinating neuropathy | 35.9 ± 26.6 | 2.7 ± 1.0<br>** | 12.9 ± 7.1<br>**** | 1.1 ± 0.9<br>n.s. |
| murine sural nerve | 77.2 | 1.9 ± 0.44<br>n.s. | 8.8 ± 1.5<br>n.s. | 1.1 ± 0.44<br>n.s. |

#### Dispersion of the paranodal junctional complex in patients with acute and severe axonal neuropathy

Next, we assessed nodo-paranodal parameters of > 180 nodes per cohort in patients classified as acute axonal, chronic axonal and chronic demyelinating neuropathy. We found a significant increase in nodo-paranodal length in all three groups compared to intact human nodes (see Figure 6 A-B). Particularly notable is that we detected apparent differences within single patients and histopathological categories: Patients with acute axonal damage showed significantly increased nodo-paranodal length compared to patients with chronic axonal polyneuropathy (15.6 vs. 12.4 μm, p = 0.002). Results closely missed significance compared to patients with demyelinating polyneuropathy (15.6 vs. 12.9 μm, p = 0.06, see Figure 6 B). Additionally, nodo-paranodal length was significantly increased in patient samples with histopathologically severe axonal loss compared to moderate (16.8 vs. 12.8, p = 0.0018) and mild axonal loss (16.8 vs. 12.9, p = 0.0022, see Figure 6 C). Interestingly, single patients showed severe and highly significant nodo-paranodal alterations, especially in the cohort of acute axonal neuropathy, whereas other patients showed nodo-paranodal lengths without significant differences to normal values (individual data are shown in Supporting Table S5). Furthermore, we compared nodo-paranodal lengths between patients fulfilling diagnostic criteria for diseases with different underlying pathophysiology: 1) Vasculitic neuropathy with primarily axonal damage and distal Wallerian degeneration due to ischemia and 2) chronic inflammatory demyelinating polyradiculoneuropathy (CIDP) affecting the internodal myelin. Nodo-paranodal length was significantly longer in patients with vasculitic neuropathy compared to CIDP (13.2 ± 6.8 vs. 12.3 ± 6.4 μm, p = 0.036). Thus, an increase in nodo-paranodal length was identified as an indicator for acute and severe axonal damage during Wallerian degeneration, and high inter-individual differences highlight possible distinct underlying pathologies and the diagnostic potential of the method. Results on further morphometric parameters and their relation to clinical information are described in the Supporting Note 4. In conclusion, the results from the high-content analysis of teased fiber samples labeled with standard immunofluorescence add to and support our findings on an ultrastructural level assessed by super resolution *d*STORM: The nodes assessed by *d*STORM were representative within the samples and diagnostic categories, and we showed that nodo-paranodal alterations occur at a severe level especially in acute and severe axonal neuropathy. By *d*STORM, we could identified the alteration of the axonal cytoskeletal ultrastructure as a possible pathogenic link to dispersion and loss of the paranodal axoglial complex in acute axonal neuropathies. A summarizing schematic display is presented in the graphical abstract (p.1).

**Fig. 6.**
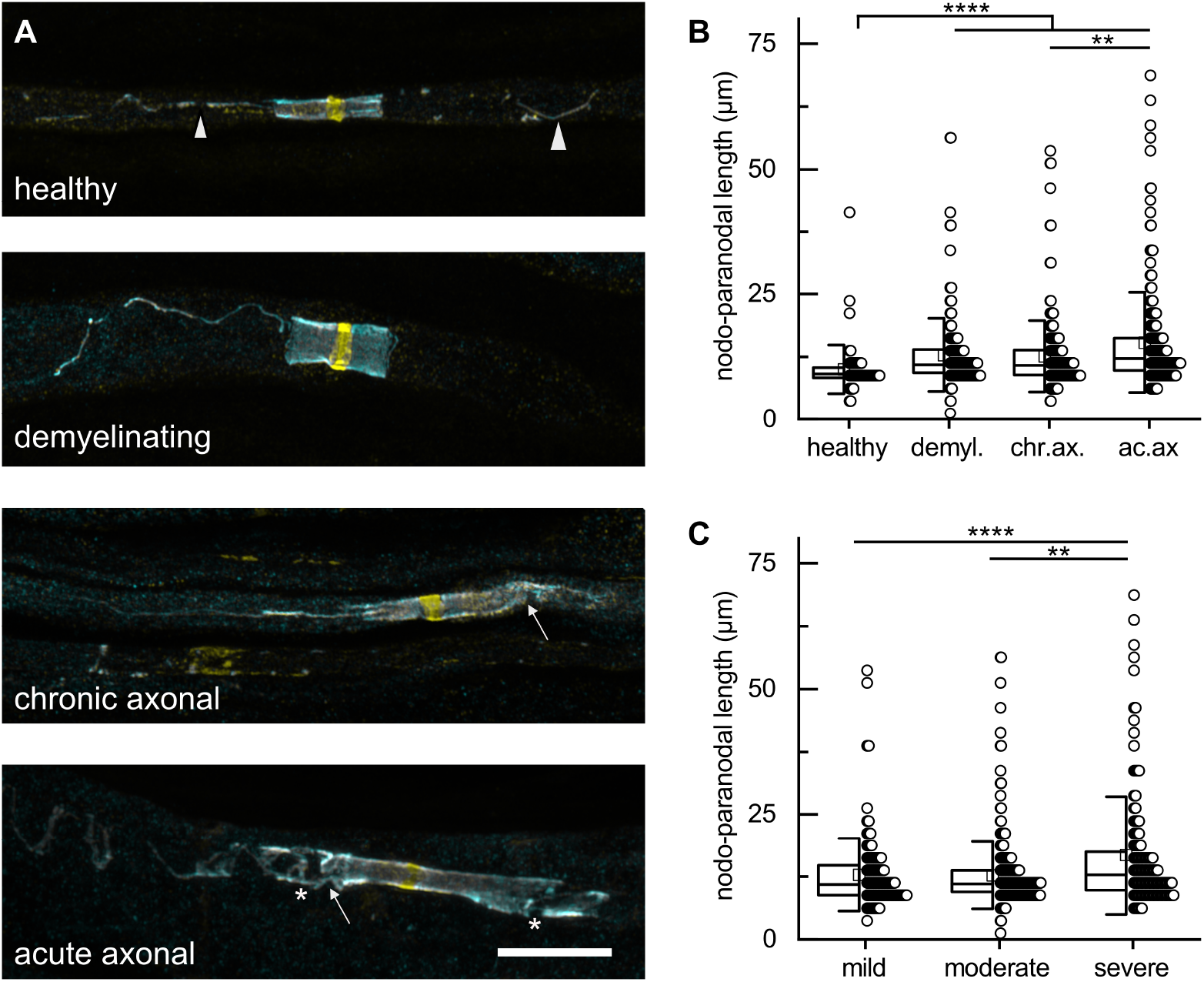
Nodo-paranodal length is elongated with patients with acute and severe axonal neuropathy. (A) High magnification photomicrographs show node and paranode morphology in human nodes of Ranvier considered as unaltered and in patients classified as demyelinating, chronic axonal and acute axonal neuropathy. In physiological nodes, paranodal Caspr-1 (cyan) and neurofascin (yellow) are dense, and the mesaxon can be distinguished clearly (arrowheads). In patients with chronic and acute axonal neuropathy, paranodal elongation with decoiling of paranodal axoglial proteins (arrows) and partial loss of staining (asterisks) is visible. Correspondingly to results of *d*STORM analysis, Caspr-1 and neurofascin colocalize at the paranode (white), even in pathologically severely altered nodes of Ranvier. Scale bar = 10μm. (B) Nodo-paranodal length (μm, y-axis) is elongated in patients with all types of polyneuropathies compared to unaltered human nodes, especially in patients with acute axonal neuropathy. (C) Nodo-paranodal length (μm, y-axis) is elongated in patients with histopathologically severe axonal loss, compared to mild and moderate axonal loss. Testing was performed using Kruskal-Wallis with Dunn’s correction for multiple testing. Significance level: * p < 0.05. ** p < 0.01, *** p < 0.001, **** p < 0.0001.

## Discussion

Here, we implement *d*STORM imaging on human tissue, map physiological axonal ultrastructure in humans and identify distinct pathologies at the node of Ranvier on a nanoscale level in polyneuropathy. We demonstrate a physiological ∼190 nm periodic arrangement of the membrane-associated cytoskeleton that was disrupted in patients with polyneuropathy, exclusively at the paranodal region. Our data indicate that cytoskeletal alterations during Wallerian degeneration might facilitate nodo-paranodal detachment. We identify nanoscale pathologies in individual patients and thereby introduce single molecule localization microscopy as a potential method to enhance diagnostics of neurological diseases.

### The ultrastructural paranodal protein arrangement is impaired in polyneuropathy

We report an increase in periodic distance of the axoglial cell adhesion molecules Caspr-1 and neurofascin-155 in patients with polyneuropathies. These proteins are components of the septate-like junctions at the paranodal region and are crucial for the integrity of the node of Ranvier and saltatory conduction (6, 9, 13). Previous studies have proposed a helical arrangement of Caspr-1 along the axon (36, 37). Here, we show a lateral increase in the periodic distance of Caspr-1 as a correlate of a lateral de-coiling of the axonal Caspr-1 helices in polyneuropathy.

### Dual-color super-resolution imaging reveals distinct pathomechanisms of paranodal disjunction

To address the origin of the decoiling of the axoglial complex, we performed dual-color colocalization studies on *d*STORM image reconstructions. Our data show that the axoglial complex of Caspr-1 and neurofascin-155 itself is robust against disjunction, even in pathologically altered paranodes. We observed either physiological paranodal axoglial arrangement, equally increased periodicity or a partial loss of both proteins in parallel. Correspondingly, previous studies using rodent models have shown a close attachment of Caspr-1 to neurofascin-155 during myelination and in demyelinating conditions (37, 38). Thus, the disruption of the nodo-paranodal architecture in classical demyelinating and axonal polyneuropathy does not occur due to a primary disjunction of the axoglial contact, which is in contrast to antibody-mediated nodo-paranodopathies (see Supporting Note 5). Importantly, our data indicate that the paranodal region constitutes an Achilles’ heel for cytoskeletal damage in axonal neuropathies. At the internode, we found no differences on internodal ß2-spectrin periodicity between humans and rodents. Our data thus are in line with previous studies that postulated that the axoskeletal ultrastructure follows a 190 nm periodicity and is conserved within species (25, 26, 30, 39).

At the paranode, on the other hand, we observed an increase in periodic arrangement of ß2-spectrin in patients with polyneuropathy compared to rodents. We exclusively studied ß2-spectrin, as other cytoskeletal proteins like F-actin and ankyrin B are also present in the paranodal glial terminal loops, and their complex 3D structure impede the analysis of axonal periodicity (14). The paranodal region might be prone to cytoskeletal alterations for two main reasons: Firstly, the axon diameter is reduced, axoskeletal transport proteins are condensed and axonal transport rates are reduced at the paranode (40, 41). Endogenous metabolites could therefore accumulate, leading to axoskeletal damage. Secondly, the node of Ranvier is less protected by the myelin sheath and thus exposed to exogenous noxious and inflammatory agents, leading to cytoskeletal alterations (42). Rodent electron microscopy studies have shown that Wallerian degeneration begins at the nodal and paranodal axon, with accumulation of cell organelles and loosening of neurofilaments (43–45). Here, we show that the paranodal axon is the initial site of Wallerian degeneration in human disease, and provide evidence for specific cytoskeletal alterations at a nanoscale and protein level in patients with axonal neuropathy as a correlate of a beginning Wallerian degeneration.

### Super-resolution imaging visualizes the link of paranodal detachment with cytoskeletal damage

Our findings imply that in axonal neuropathies, the paranodal cytoskeletal damage might be associated with the nodo-paranodal detachment. Previous studies including *d*STORM data suggest that a2/ß2-spectrin serves as an intra-axonal barrier, is crucial for the clustering of sodium channels at the node, for maintaining the periodicity of the membrane-associated cytoskeleton, and for axon survival (29, 46, 47). Here, we show that along with the alteration of the axonal cytoskeleton, the colocalization of Caspr-1 and its axonal anchor is severely disrupted. This finding implies that the alteration of cytoskeletal structure proteins might lead to a lateral decoiling of the Caspr-1 helices along the axon. This is in line with reports of a paranodal disorganization and decreased expression levels of Caspr-1 in protein 4.1B murine knockout models (32, 48, 49). As we can show that Caspr-1 and glial Neurofascin-155 attach strongly even if pathologically altered, we propose that Caspr-1 decoiling and loss might cause a broadening and detachment of the paranodal loops from the axon, with paranodal lengthening as a consequence. Correspondingly, we found a correlation of nodo-paranodal length to histopathological severity and acuteness of axonal loss. In line with these findings, early ultrastructural studies reported terminal loop alterations after cytoskeletal changes during Wallerian degeneration (43, 44). Paranodal detachment was previously reported in other neuropathies classified as axonal, such as diabetic neuropathy or chronic idiopathic axonal neuropathy (50, 51). Interestingly, patients of our cohort with primarily axonal pathology such as vasculitic neuropathy showed an increased nodo-paranodal length compared to CIDP, a disease characterized by internodal demyelination. Previously, Caspr-1 staining was proposed as a diagnostic marker to distinguish chronic idiopathic axonal neuropathy from CIDP, in line with our findings (51). Still, the conditional murine knockout of single axoskeletal molecules, even ß2-spectrin, does not lead to a complete detachment of Caspr-1, previously shown by using diffraction-limited microscopy (7, 46). Therefore, we suggest that a paranodal disjunction in axonal neuropathy occurs in case of a severe disruption of the axonal cytoskeleton involving several structure proteins at a time, and can better be observed using superresolution imaging. Further studies are needed to address various cytoskeletal proteins, especially investigating the role of protein 4.1B (31, 48, 49), which was not included in the study due to a lack of a commercial antibody apt for super resolution microscopy. In summary, our data indicate that a paranodal elongation and disruption may rather be considered a secondary effect of axonal damage than a primarily demyelinating feature in axonal neuropathy. Via a combination of *d*STORM and high content confocal imaging, we could show that acute Wallerian degeneration with consecutive cytoskeletal damage could lead to dispersion of proteins of the axoglial complex and paranodal detachment.

### Advanced high- and super-resolution microscopy pushes the boundaries of diagnostic assessment of nerve biopsies in polyneuropathies

Using a high-content confocal imaging approach, we were not only able to confirm our histopathological findings on an ultra-structural level in high sample sizes, but also provide a protocol of possible future use in the diagnostic work-up of polyneuropathies. Single teased fiber analysis, electron microscopy or longitudinal sections are well-established methods to examine the node of Ranvier, additionally to gold-standard cross-sectional analysis (52–55). Still, these methods can be time-consuming, require high expertise due to non-standardized assessment, depend on rater-dependent accuracy and partly do not allow to specifically study single proteins of interest (52–54, 56). Here, we introduce a different protocol including fiber bundle teasing that can be performed within 60 minutes, followed by triple immunofluorescence staining and automated confocal high-content image acquisition. This method yields the opportunity to study numerous nodes of Ranvier at high magnification, with easy navigation within the sample due to automated identification of regions of interest, and could reduce selection bias by the examiner. Furthermore, we applied basic deep learning algorithms on teased fiber preparations to assess nodal count and nodo-paranodal length, which was previously only reported in rodent samples and human cross-sectional tissue (see Supporting Note 3). Further development of these methods in future studies may set the basis for modern image analysis and quantification approaches to enhance histopathologic assessment of polyneuropathy. Furthermore, the imaging routine implemented in this study could in the future be transferred to less invasive diagnostic samples, such as skin punch biopsies, or to samples beyond neuropathies. We report standardized normal values of human adult sensory nerve nodo-paranodal length that had previously only been defined in children (57), and identify the node of Ranvier density as a surrogate marker for axonal loss in teased fiber preparations. Thus, we provide a method to assess both severity of axonal neuropathy and node of Ranvier pathology using one single application. Larger studies with homogenous cohorts need to confirm if nodo-paranodal morphometrics and ultrastructural assessment via *d*STORM can serve as diagnostic markers and assess their sensitivity, specificity and practicability in routine assessment.

In conclusion, our data contribute to the understanding of underlying node of Ranvier pathologies of peripheral neuropathies on a nanoscale level, and highlight the potential of advanced high and super-resolution microscopy for both diagnostics and pathophysiological studies. Our findings could pave the way for introducing new imaging techniques and super resolution microscopy into histopathologic diagnostic assessment of patients with neurologic conditions.

## Data Availability

All data produced in the present study are available upon reasonable request to the authors.

https://doi.org/10.5281/zenodo.6387771

## Data availability

Data as described above are available here: https://doi.org/10.5281/zenodo.6387771. Further raw data including original photomicrographs and simulations will be available upon request by any qualified researcher.

## Study approval

The study was approved by the institutional ethical committee (approval number 277/13 and 15/19). All subjects gave written informed consent prior to participation.

## ACKNOWLEDGEMENTS

The study was supported by a grant of the Interdisciplinary Center of Clinical Research (IZKF) of the Medical Faculty of Würzburg to K.D. and K.G.H (F-N-439). J.L. was supported by a scholarship of the Graduate School of Life Sciences (GSLS) of the University of Würzburg. H.S.H. was supported by the Rudolf-Virchow-Center of the University of Würzburg. L.A. and K.D. hold research fellowships by the Interdisciplinary Center of Clinical Research (IZKF) of the Medical Faculty of Würzburg. We thank Antonia Kohl and Barbara Reuter for excellent technical assistance and A. Balakrishnan and paraphrasis.de for proof-reading. We thank the patients and animals who contributed to the study.

## AUTHOR CONTRIBUTIONS

K.G.H., K.D., L.A. and H.S.H conceived and designed the experiments. H.S.H. developed the image-correlation analysis routine. J.L. and C.K. performed the sample preparation, imaging and data analysis. J.S. and K.H. performed the image segmentation and analysis of the AiryScan data. K.D. assessed histopathological features. L.A. assessed clinical data and analyzed clinical, histopathological and super resolution microscopy data. L.A., H.S.H, and J.L. wrote the first draft of the manuscript. C.S., K.G.H., and K.D. revised the manuscript for scientific content. All authors read and approved the final manuscript.

## Online Methods

### Patients, clinical and histopathological diagnosis

We prospectively included n = 17 patients with peripheral neuropathies who had undergone a sensory nerve biopsy during the routine diagnostic workup process at the University hospital of Würzburg between July 2018 and April 2021. Clinical and histopathological data were assessed as described in the Supporting Note 1 and are shown in Supporting Table S1 and S2. The study was performed in line with the principles of the Declaration of Helsinki and was approved by the institutional ethical committee (approval number 277/13 and 15/19). All subjects gave written informed consent prior to participation.

### Teased fiber preparation

Murine teased fibers we prepared as previously described (58). In brief, adult C57/Bl6 mice were sacrificed by cervical dislocation. Sciatic and sural nerve were dissected, fixed for 10 minutes in 4% PFA, and washed in 1M phosphate buffer. Human sensory nerve preparations (n = 16 sural nerve, n = 1 superficial radial nerve) were obtained from leftovers of routine nerve biopsies of patients with polyneuropathies undergoing a diagnostic workup process at the University Hospital of Würzburg. The nerve was cut into 5-mm slices with a scalpel, fixed and washed as described above. Epineural connective tissue and the perineurium were removed with fine forceps. The remaining nerve fascicle was including endoneurial connective tissue was teased with fine forceps in 0.5M phosphate buffer, so that single axons and /or bundles were separated. For *d*STORM microscopy, the fibers were teased on ultra-cleaned, PDL-coated 24-mm glass coverslips. For confocal microscopy, the fibers were teased on Airyscan Superfrost Slides. Slides and coverslips were dried overnight and stored at -20°C until use. The teasing process was performed by a technician or investigator and two or less attempts on learning the teasing process resulted in sufficient results. The whole procedure of human sural nerve fixation, preparation and teasing took a maximum of 60 minutes or less.

### Simulated Datasets

Simulations of super-resolution microscopy datasets were performed with the SuReSim software package (59). The periodic ground truth structure is available on Zenodo (see data sharing statement) and the simulation parameters are summarized in Supplemental Table S6). The localization text-files were transformed into ThunderSTORM (60) compatible csv-files and super-resolution images with a custom MatLab script. Simulated datasets were analyzed the same way as the experimental datasets (see below).

### Immunostaining for *d*STORM and confocal microscopy

The thawed teased fibers were fixed for 20 minutes in 100% Acetone at -20 °C. After washing with Phosphate-buffered saline (PBS) and permeabilization with 2% Triton-X for 30 min, they were blocked in PBS containing 1mg/ml Biotin-BSA, 4% BSA and 0,5% Triton-X for 60 min. Dual-color immunolabeling was performed by overnight incubation at + 4 °C with the primary antibody diluted in PBS with 0.5% Triton-X and 1% BSA (see Supplemental Table S7). For *d*STORM imaging, fiducial markers were immobilized on the surface by an additional 20 min incubation step with 0.1 mg/ml Neutravidin (Thermo Scientific™, 31000) and three PBS washing steps right before the initiation of the secondary antibody incubation. During the secondary antibody incubation 35 pM biotinylated gold nanorods (Nanopartz C12-25-650-TB) were added. After secondary antibody incubation for 1h at room temperature, slides and coverslips were washed and *d*STORM coverslips were stored PBS. For Airyscan imaging, the slides were embedded in Vectashield antifade mounting medium (Vector laboratories, VEC-H-1000).

### *d*STORM Fluorescence image acquisition

Dual-color *d*STORM microscopy was performed with an inverted light microscope (Zeiss Observer Z.1, Carl Zeiss AG) customized for SMLM (Supplemental Table S8). The *d*STORM blinking buffer contained 100 mM MEA (Sigma, M6500), 20 mM D-glucose (Sigma, G7528), 0.55 mg ml-1 gluco-oxidase (Roth, 60281), 0.011 mg ml-1 catalase (Sigma, C1345) in PBS. The pH was adjusted to pH 7.9 with 5M KOH (Sigma, 30603). After applying the *d*STORM buffer, imaging was performed with the acquisition parameters summarized in Supplemental Table S9. For color channel alignment, calibration images with 100 nm TetraSpeck microspheres (ThermoFisher, T7279) immobilized on glass coverslips were recorded simultaneously in both channels.

### *d*STORM image reconstruction

*d*STORM images were reconstructed with the ImageJ plugin ThunderSTORM (60, 61) as specified in Supplemental Table S9). Based on the filtered localization data, a super-resolved image with a pixel size of 10.6 nm was reconstructed by Gaussian rendering or as a scatter plot. For the channel alignment, the TetraSpeck images were localized and a super-resolved image with a pixel size of 10.6 nm was reconstructed for each channel. Based on these calibration images, a transformation matrix of the A532 channel onto the A647 was calculated with the ImageJ plugin bunwarpJ (62). This transformation matrix was directly applied to the reconstructed super-resolved images or to the localization data itself with a MatLab script.

### *d*STORM image autocorrelation analysis

The image autocorrelation analysis was performed based on the localization data with a MatLab routine (Scripts and manuals are available on Zenodo, see Data sharing statement). With the routine, several regions of interest (ROIs) within the paranodal and internodal region were selected manually in the super-resolved reconstruction of the respective node (width = 600 nm; lengths along the axon were manually chosen by selecting a start and end point of the centerline). Based on localization data contained in each ROI, a super-resolved image was reconstructed after rotating the ROI to align the image along the x-axis. The rotation angle was varied within a ± 20° range by 1°-steps to align the repetitive line structure perpendicularly to the x-axis. The resulting reconstructed images were autocorrelated in 2D with the Normxcorr2_general.m function (63). The optimal rotation angle was selected based on a maximizing of the autocorrelation along the center y-axis. MatLab output correlation images (pixelsize 2 nm) were analyzed in FIJI (64) with line plots averaged over a width of 150 pixels. Plots were imported into OriginPro (Origin2021b, OriginLab Corp.). Gauss distribution curves were fitted and the peak to peak distances of those distribution curves were calculated.

### *d*STORM image colocalization analysis

To estimate the image resolution the raw time series image stacks were split into even and colocalization analysis on transformed dualcolor *d*STORM datasets was performed with a tessellation-based approach (21). Paranodal regions were defined by Caspr-1 staining, each representing a region of interest. Herewithin, Manders’ A & B correlation coefficients of all channels were calculated to assess colocalization (34, 35). The pan-neurofascin antibody used in this study stained both glial neurofascin-155 isoform at the paranode and neuronal neurofascin-186 isoform at the node, which have different localizations and functions (65, 66). In this study, we exclusively studied neurofascin-155 and its interaction with Caspr-1, therefore we exclusively studied the paranodal region. The nodal region within the pan-neurofascin staining was not considered in the analysis. The colocalization data was analyzed using OriginPro.

### Large field of view confocal imaging and assessment

High-content 3D three-color confocal microscopy at a 20-fold magnification including stacks and tiles was performed with a Zeiss LSM 980 with Airyscan 2 (Carl Zeiss AG) using Zen blue 3.3 software (Carl Zeiss AG) for Airyscan 3D processing and stitching of tiles. Acquisition and image processing parameters are listed in Supporting Table S9. To count and locate nodes of Ranvier within the sample and to determine the node of Ranvier density within the sample, we performed and compared either manual counting combined with background segmentation using FIJI, semi-automated threshold-based segmentation considering specific anti-pan-neurofascin staining using Imaris software (v.9.9.0, Oxford instruments, Abingdon, UK) or fully automated image segmentation using deep learning (see Supporting Notes 2 and 3). To assess normal values for nodo-paranodal morphometrics, we measured n = 69 morphologically intact human nodes of Ranvier from n = 4 patients of our cohort with mild neuropathy and only minor pathological alterations in the semithin sections (see Supporting Table S5). Healthy nodes were identified by 1) two opposing and symmetric Caspr-1 blocks, 2) homogeneous staining and 3) rectangular form. A blinded investigator performed manual measurements by defining linear ROIs using FIJI. We assessed 1) the maximal extension of two opposing Caspr-1 blocks including the nodal gap as an indicator of nodo-paranodal length, 2) the minimal gap between two Caspr-1 blocks as an indicator of the nodal length and 3) the maximal Caspr-1 diameter as an indicator of the paranodal axonal diameter (see Figure 5 A-B and the Movie M1). N = 40 randomly selected nodes of Ranvier were measured in each patient sample of n = 9 patients. In n = 7 patients, the maximal number of nodes per field of view was < 40 nodes, so all visible nodes within the field of view were assessed (n = 22 - 39 nodes, see S5). The manual analysis was compared to automated measurements of nodo-paranodal length. Accuracy of deep learning or threshold based image segmentation did not reach as exact results as manual measurement (see Supporting Note 3 and Supporting Fig. S4).

### Statistics

We assessed statistical significance with OriginPro Version 2021b SR2 9.8.5.212 (OriginLab Corp.) and Prism V9.3.0 (GraphPad Software, San Diego, CA). Testing for normality distribution was performed using d’Agostino Pearson test and Shapiro-Wilk test. The further specific tests used in the analysis are described in the respective figure and table legends. Results with p-values less than 0.05 were considered significant.

## Supplementary Material

**Supplorting Notes 1-5**.

**Supplementary Tables 1-9**.

**Supplementary Figures 1-4**.

**Supplementary Movie 1**.

**Supplementary Bibliography**.

## Supporting Notes

### Supplementary Note 1: Assessment of patients’ clinical and histopathological data

Demographic and clinical data of the n = 17 patients including laboratory results, nerve conduction studies and suspected clinical diagnosis were assessed retrospectively from the patients’ records. HbA1c values were assessed at the time point of the nerve biopsy and diabetes mellitus was classified according to WHO criteria. Diagnostic criteria for chronic inflammatory demyelinating polyradiculoneuropathy (CIDP) (1), POEMS syndrome (2) and vasculitic neuropathy (3) were assessed in patients with the respective clinical diagnosis. Clinical disease severity respective to motor function was assessed retrospectively by applying standardized clinical scores: The Overall disability sum Score (ODSS) (4) and the Medical Research Council (MRC) Sum Score for levels of paresis (5). The histopathological diagnosis was assessed in each biopsy by a trained and blinded researcher reevaluating the routine staining including semithin sections with toluidine blue staining, cryosection including H&E staining and immunohistochemistry for T-cell marker anti-CD4 and paraffin-embedded sections including H&E, Congo red and Elastica van Gieson staining and immunohistochemistry for macrophages using anti-CD68 and lymphocytes using anti-CD8 according to standard protocols as previously described (6). Onion bulb structures, regeneration clusters, thinly myelinated fibers and Wallerian degeneration were counted per fascicle in each patient. Axonal loss was quantified in a semi-quantitative manner (mild, moderate, severe) and large fibers per fascicle were counted and the area of the fascicle was measured using FIJI (7, 8) to calculate the fiber density (unit: fibers/mm^2^).

### Supplementary Note 2: Implementation of automated image analysis methods and deep learning strategies to assess nodes of Ranvier in teased fiber preparations

#### A. Threshold based segmentation

To count and locate nodes of Ranvier, we performed semi-automated threshold based image segmentation of the specific anti-pan-neurofascin staining using Imaris (v9.9.0, Oxford instruments, Abingdon, UK). Imaging smoothing was set to 2 times pixelsize (0.15 μm). The intensity threshold was set manually to values between 500 - 1400 to include stained nodes of Ranvier individually for every image. Results were filtered by object size (5000 voxel). Analysis was performed in n = 20 high-content images of murine sciatic nerves and human samples each. For comparison and accuracy assessment, the number of nodes per sample was counted manually using FIJI by two independent and blinded investigators. To determine the node of Ranvier density within the sample, the total tissue volume was calculated based on unspecific background binding of the anti-pan-neurofascin antibody using a custom FIJI script (available on Zenodo, see data sharing statement). Images were filtered with a 3D Gaussian blur and then classified into “tissue” and “non tissue” by a low static threshold. Morphometrics of the nodes of Ranvier were assessed and compared via two methods: Firstly, nodo-paranodal length was calculated from Imaris ellipsoid axes of pan-neurofascin staining in n = 20 slides of murine and human samples (see above and Supporting Fig. S4). Secondly, morphometrics were assessed on the same slides in a blinded manner by FIJI defining linear ROIs as indicated in Figure 5.

#### B. Automated image segmentation using deep learning

Fully automated image segmentation of specific anti-pan-neurofascin stained nodes of Ranvier with an artificial neural network was performed using custom python scripts utilizing ResNet-101 retrieved from the Pytorch library (9, 10). First, individual z-slices of high content images were split into smaller tiles (224*224 px) and individually downscaled with a min/max scaling. Image tiles were then converted to RGB and transformed with the network’s default transformation functions. Training for patient high content image segmentation was performed using 40 randomly selected image slices and 100.000 training iterations with batches of 50 tiles per iteration. Loss was calculated via cross entropy, weights were optimized using the Adam optimizer and a learning rate of 1e-6. To minimize network habituation to the training data, three layers of randomization were used for every iteration: A random image was opened, the seed point for tile creation was randomized and the tiles included in the batch were randomly chosen. The final prediction of the pixel classification was calculated twice with different seed points for tile creation. Only pixel detected in both classifications were included to minimize tile border effects. Analysis of the pixel classification data was performed using the skimage image library. Objects were labeled and object statistics (area, position, axis length, intensities of the original image) were calculated with skimage built-in functions on a max-projection.

### Supplementary Note 3: Implementing deep learning strategies and automated image analysis methods on teased fiber assessment

To achieve accurate, unbiased counting and morphometrics and to seek ways to enhance diagnostic assessment of nerve biopsies in the future, we implemented automated image analysis methods on large field-of-view photomicrographs of teased fiber preparations (see above and Supporting Fig. S4 A-C). Counting accuracy of the node of Ranvier density was compared to manual assessment. As a result, deep learning (DL) based median counting accuracy in murine datasets was 0.99 (0.75 - 1.15) and 1.21 (0.09 - 4.43) in human datasets. The semi-automated, segmentation-based approach failed toreach the same accuracy, with a median of 1.21 (0.97 - 2.42) for murine datasets and 2.83 (0.38 - 10.0) in human datasets, with significant differences compared to the DL based strategy (see Supporting Fig. S4 D). The nodal count correlated between manual assessment and DL based assessment when compared in single samples both for murine and human datasets (Spearman r = 0.91, p < 0.0001 and r = 0.87, p < 0.0001). Recognition using segmentation had a reduced counting accuracy with a significantly increased variance (mouse: F = 0.033, p < 0.05 human: F = 0.033, p < 0.0001), possibly due to the heterogeneity of pathological alterations. Thus, we could show that DL based routines can accurately count nodes of Ranvier in teased fiber bundles in a fully automated manner, and are superior to segmentation-based approaches. Furthermore, we seeked to implement automated morphometrical analysis including assessment of nodo-paranodal length via DL and threshold based approaches. Therefore, we assessed nodo-paranodal length from the object statistics, which were extracted from the detected nodes of Ranvier by DL and threshold based image segmentation, and compared them to the manually assessed measurements as described above. In both DL and threshold based analysis, the results differed significantly between automated and manual analysis both in murine samples (median nodo-paranodal length by manual assessment: 8.6 ± 1.5 vs. DL based measurement 7.2 ± 2.7 p = 0.02, vs. threshold based measurement 6.9 ± 4.2 p < 0.0001) and in human samples (median nodo-paranodal length per manual assessment: 10.9 ± 7.0 vs. DL based measurement 7.8 ± 4.2 p < 0.001, vs. threshold based measurement 6.1 ± 31.0 p < 0.0001). Thus, automated image analysis did not yet reach the same accuracy as manual measurement by two blinded, independent investigators, especially in pathologically altered samples (see Supporting Fig. S4E). In conclusion, we can thus show that deep learning based assessment can add to the diagnostic assessment of teased fiber analysis. In previous studies, segmentation techniques and deep learning algorithms have not only been applied on super resolution data, but also on rodent and porcine peripheral nerve and human healthy CNS cross sections (11–15). Most recently, super resolution microscopy and image segmentation was successfully implemented on healthy human peripheral nerve cross sections (16). Our data provide a new application for these new techniques. Better training sets and new DL algorithms may also allow for DL to be used for morphometric assessment in the future, and thus enhance diagnostic means. As an outlook, the DL based methodology introduced in this study paves the way to navigate within a sample and to establish automated target recognition in large and dense samples, with following dSTORM imaging of regions of interest. Thereby, nanoscale pathology could be assessed in a fully automated manner, and thus be translated to routine diagnostics. Future studies need to address and further investigate these new approaches.

### Supplementary Note 4: Supporting Note S2: Morphometric analysis of patients’ teased fiber preparations using high-content confocal imaging

We assessed nodo-paranodal length, paranodal diameter and nodal length as described in detail in the methods in the main manuscript. Results on nodo-paranodal length are described in detail in the main manuscript. The nodal length did not differ between patient cohorts with acute, chronic axonal and demyelinating neuropathy compared to intact nodes, nor between patient cohorts with mild, moderate or severe axonal loss (see Table 1). Still, we found significant nodal elongations in single patients who were clinically severely affected (see Supporting Table S3). In the whole cohort, the nodal length correlated to clinical motor function scores (mean nodal length vs. Overall disability sum score: Pearson r = 0.59, p = 0.01; mean nodal length vs. Medical Research Council Sum Score: Pearson r = -0.83, p < 0.0001), but not to histopathological severity of axonal loss. Most patients with axonal neuropathies showed normal paranodal axonal diameters (see Table 1 and Supporting Table S5). On the other hand, the paranodal diameter in patients with chronic demyelinating polyneuropathy was 2.7 μm and thus significantly increased compared to intact nodes, and compared to patients with acute and chronic axonal neuropathy (p < 0.0001). Thus, paranodal swelling was found to be prominent in patients with demyelinating neuropathies. We did not detect any correlation of clinical or histopathological disease severity and axonal diameter. In all analyses, age was considered as a possible confounder, but did not correlate to any of the measured parameters. In conclusion, the morphometric data on nodal length and diameter help to discriminate single patients regarding clinical disease severity and type of neuropathy. Despite a small sample size, we identify paranodal swelling as exclusive for demyelinating disease, as previously described in avian models of demyelination and in patients with multiple sclerosis (17, 18). Furthermore, we find nodal elongation in single patients as a possible indicator of clinically severe demyelinating disease, corresponding to previous reports on nodal elongations in patients with in CIDP (19). Thus, these parameters may be valuable in histopathological diagnostic assessment of patients with neuropathies of unknown etiology, but need to be validated in larger studies with homogenous cohorts. As a limitation, we cannot address yet to what extent internodal and nodo-paranodal changes are physiological and occur with age in the elderly due to a lack of human healthy controls due to ethical reasons, as post-mortem autopsy material as a control might not be suited due to immediate and early post-mortem cytoskeletal and structural alterations (20). Still, we did not observe a correlation of age to nodo-paranodal parameters, arguing against age being a confounder in the analysis.

### Supplementary Note 5: Paranodal detachment in axonal neuropathy differs from autoimmune nodo-paranodopathy and demyelinating neuropathy

The pathomechanism of nodo-paranodal disruption in axonal neuropathies we propose within this study is different to previously suggested mechanisms of nodo–paranododal disruption in autoantibody-mediated nodo-paranodopathy. This rare disease is an autoimmune neuropathy associated with pathogenic autoantibodies against proteins located at the node of Ranvier, which attack proteins of the axoglial junction. As a consequence, a primary axoglial detachment induced by the autoantibodies has been proposed, either by directly inhibiting the interaction of axonal and glial binding partners or by altered myelin turnover (21–25). Due to its characteristic clinical and pathophysiological features, autoimmune neuropathy has been proposed as a new sub-entity within immune-mediated neuropathies (1, 26). However, our cohort does not include patients with this rare disease, as today, antibodies serve as a biomarker and reduce the need for invasive diagnostic procedures. Additionally, nodal elongations measured by the gap of myelin basic protein have also been described in skin biopsies of patients with chronic inflammatory demyelinating neuropathy (19, 27, 28). In this study, we found paranodal elongations measured by Caspr-1 staining in patients with acute axonal neuropathy to be significantly longer than in CIDP. Still, our CIDP cohort is small and not typical, as diagnosis of CIDP usually does not need nerve biopsy and is only performed in unclear cases. We thus cannot directly compare these possible mechanisms of axoglial disjunction. Still, we hypothesize that in axonal neuropathies, the axoglial complex remains intact, but detaches from its cytoskeletal anchor, whereas autoimmune nodopathies lead to a axoglial detachment (24, 25, 29–31). The methods described in this study yield the opportunity to investigate and directly compare different mechanisms of paranodal damage on a nanostructural scale in future studies, e.g. including cell-culture-based assays.

## Supplementary Tables

**Table S1.**
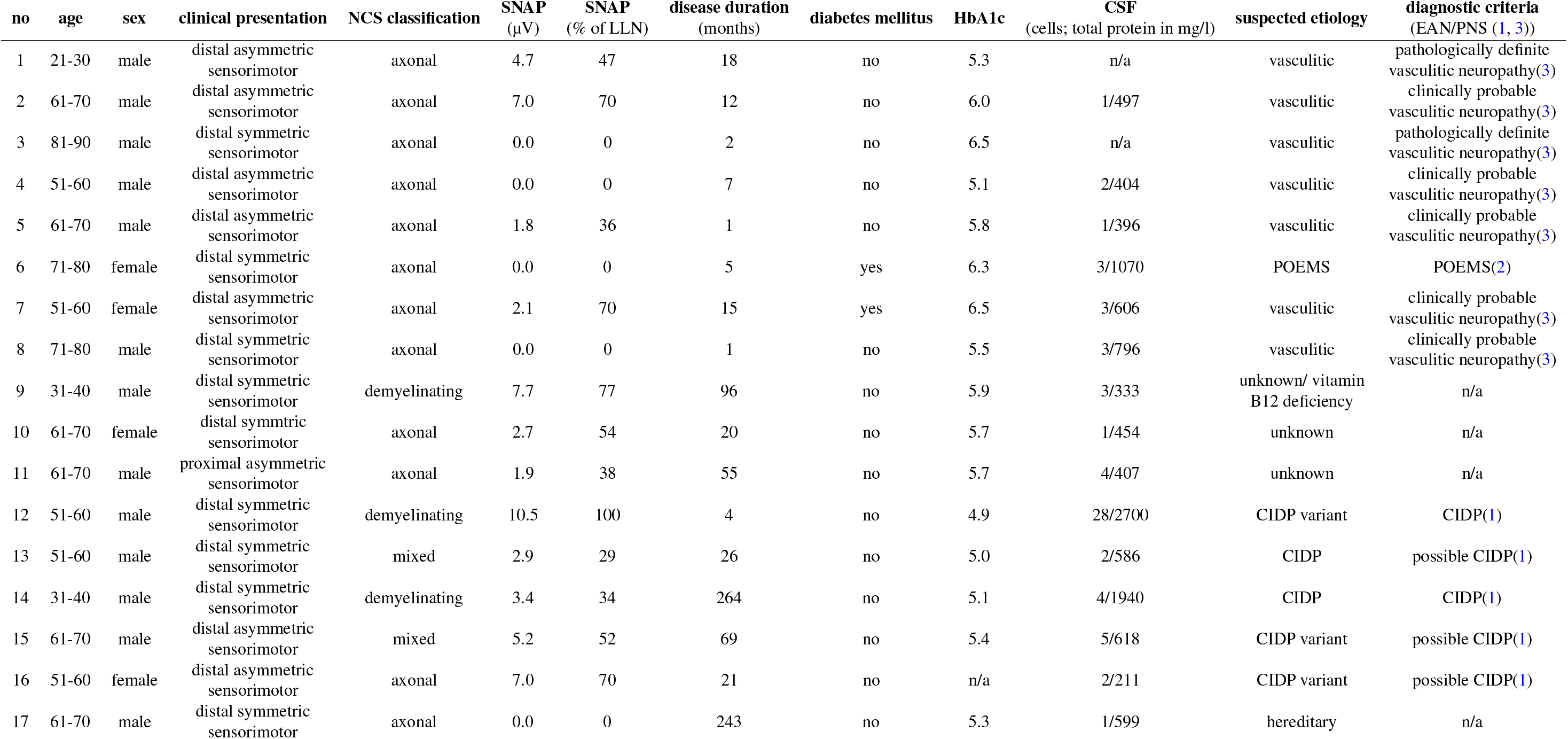
Demographic data and clinical of the cohort of patients with polyneuropathy. Abbreviations: CIDP = chronic inflammatory demyelinating polyradiculoneuropathy, CSF = cerebrospinal fluid, n/a = not applicable or not assessed, LLN = lower limit of normal, NCS = nerve conduction studies, no = number, POEMS = polyneuropathy, organomegaly, endocrinopathy, myeloma protein, and skin changes (osteosclerotic myeloma), SNAP = Sensory nerve action potential.

**Table S2.** Histopathologic parameters and classification of patients with polyneuropathy. In each patient, semithin cross sections of sensory nerve biopsy (patient 7: superficial radial nerve, other patients: sural nerve) with toluidine blue staining were evaluated for histopathological classification. Total counts of onion bulb figures, thinly myelinated fibers, Wallerian degeneration and large fibers are displayed for two fascicles per patient. Fascicle area was measured using FIJI. Fiber density was calculated as n(large fibers) / mm^³^ and is displayed both for the single fascicles and as the mean fiber density. Furthermore, immunostainings were evaluated as described in the methods to check for signs of inflammation and amyloid deposition, and if positive, are reported above. Histopathologic classification including semiquantitative assessment of axonal loss, acuteness of disease and type (axonal or demyelinating) was performed by a trained and blinded researcher. Abbreviations: no = number.

| no | onion<br>bulbs | regenerating<br>clusters | thinly<br>myelinated<br>fibers | Wallerian<br>degeneration | large<br>fibers | fascicle area<br>($\mu\text{m}^2$) | fiber density<br>(large fibers/$\text{mm}^2$) | mean<br>fiber<br>density | signs of inflammation/vessel pathology | severity<br>axonal<br>loss | acuteness | type |
| --- | --- | --- | --- | --- | --- | --- | --- | --- | --- | --- | --- | --- |
| 1 | 0/0 | 0/0 | 2/2 | 2/1 | 297/129 | 131469/114096 | 2259/1131 | 1695 | perivascular leukocytes (including the vessel wall),<br>fragmentation of lamina elastica interna, necrosis | moderate | acute | axonal |
| 2 | 0/0 | 2/1 | 3/0 | 4/5 | 222/132 | 118621/68965 | 1872/1914 | 1893 | perivascular leukocytes (including the vessel wall) | moderate | acute | axonal |
| 3 | 0/0 | 0/2 | 0/0 | 16/6 | 50/141 | 130332/117842 | 384/1197 | 790 | perivascular leukocytes (including the vessel wall),<br>fragmentation of internal elastic lamina | moderate | acute | axonal |
| 4 | 0/0 | 2/0 | 0/0 | 9/6 | 10/11 | 114664 / 98878 | 87 / 111 | 99 | perivascular leukocytes (including the vessel wall),<br>endoneurial macrophages | severe | acute | axonal |
| 5 | 0/0 | 0/0 | 0/0 | 32/42 | 32/42 | 103204/72460 | 230/745 | 488 | perivascular leukocytes (including the vessel wall),<br>endoneurial macrophages | severe | acute | axonal |
| 6 | 0/0 | 4/0 | 4/2 | 3/11 | 38/29 | 105348/54121 | 361/546 | 453 | perivascular leukocytes | severe | acute | axonal |
| 7 | 0/0 | 2/2 | 3/1 | 0/2 | 67/410 | 57923/107453 | 1157/3816 | 2486 | perivascular leukocytes, endoneurial macrophages | mild | chronic | axonal |
| 8 | 5/1 | 9/6 | 4/1 | 0/0 | 71/161 | 58634/103384 | 1211/1557 | 1384 | perivascular leukocytes, fragmentation of<br>elastic lamina | moderate | chronic | axonal |
| 9 | 3/3 | 3/2 | 3/3 | 0/0 | 121/98 | 113839/55362 | 1063/1770 | 1417 | none | moderate | chronic | axonal |
| 10 | 0/0 | 7/5 | 18/12 | 0/0 | 185/143 | 85716/51282 | 2158/2788 | 2473 | none | moderate | chronic | axonal |
| 11 | 0/0 | 3/4 | 4/2 | 3/3 | 191/134 | 62462/42427 | 3058/3158 | 3108 | endoneurial macrophages | mild | chronic | axonal |
| 12 | 3/0 | 8/5 | 5/2 | 0/0 | 552/218 | 104192/75243 | 5298/2897 | 4098 | perineurial leukocytes | mild | chronic | axonal |
| 13 | 5/0 | 2/0 | 35/3 | 2/1 | 98/24 | 66966/19482 | 1463/1232 | 1348 | perivascular leukocytes, perivascular and<br>endoneurial macrophages | severe | chronic | demyelinating |
| 14 | 3/6 | 7/6 | 18/3 | 0/0 | 205/196 | 104881/115955 | 1955/1690 | 1822 | endoneurial and perivascular leukocytes,<br>endoneurial macrophages | moderate | chronic | demyelinating |
| 15 | 2/1 | 4/6 | 29/41 | 0/1 | 125/142 | 52967/194474 | 2360/1359 | 1860 | none | moderate | chronic | demyelinating |
| 16 | 2/2 | 0/1 | 12/7 | 0/1 | 215/155 | 87379/54731 | 2461/2832 | 2646 | endoneurial and perivascular macrophages | mild | chronic | demyelinating |
| 17 | 2 | 6/2 | 24/6 | 0/0 | 110/95 | 78121/55787 | 1408/1703 | 1555 | none | moderate | chronic | demyelinating |

**Table S3.** Histopathological and clinical differences between groups of histopathological classification. For histopathological classification, patients were classified into histopathological 3A) type (axonal and demyelinating neuropathy) by counting signs of demyelination such as onion bulb figures and thinly myelinated fibers/fascicle. Furthermore, 3B) histopathological acuity was determined by counting signs of de- and regeneration and correlated to clinical disease duration. 3C) histopathological severity was rated semiquantitatively in mild, moderate and severe and assessed quantitatively by counting the large fiber density/mm^2^. Electrophysiological parameters (SNAP) were assessed and compared in the three histopathological severity groups. The last column shows results of statistical testing between the groups using Kruskal-Wallis test with Dunn’s correction for multiple testing if necessary (adjusted p-values are shown). Abbreviations: LLN = lower limit of normal, SNAP = Sensory nerve action potential.

| 3A) type | axonal<br>(n=12) | demyelinating<br>(n=5) | p-value |  |
| --- | --- | --- | --- | --- |
| median count of<br>onion bulbs/fascicle | 0 | 2 | 0.0047 |  |
| median count of<br>thinly myelinated fibers/fascicle | 2.3 | 15 | 0.0015 |  |
| 3B) acuteness | acute<br>(n=6) | chronic<br>(n=12) | p-value |  |
| median Wallerian degeneration<br>figures/fascicle | 7.3 | 0.0 | 0.0003 |  |
| median count of regenerating<br>clusters / fascicle | 1.0 | 4.0 | 0.0076 |  |
| median disease duration (months) | 6 | 42 | 0.021 |  |
| 3C) severity | mild<br>(n=4) | moderate<br>(n=8) | sever<br>(n=4) | p-value |
| mean large fiber density<br>(1/mm<sup>2</sup>) | 3085 ± 725 | 1654 ± 459 | 597 ± 531 | $P_{mild\,vs.\,sever} < 0.0001$<br>$P_{moderate\,vs.\,sever} = 0.0176$ |
| SNAP (% of LLN) | 69.6 ± 25.32 | 37.11 ± 30.54 | 16.25 ± 18.98 | $P_{mild\,vs.\,sever} < 0.04$<br>$P_{moderate\,vs.\,sever} = 0.4$ |

**Table S4.**
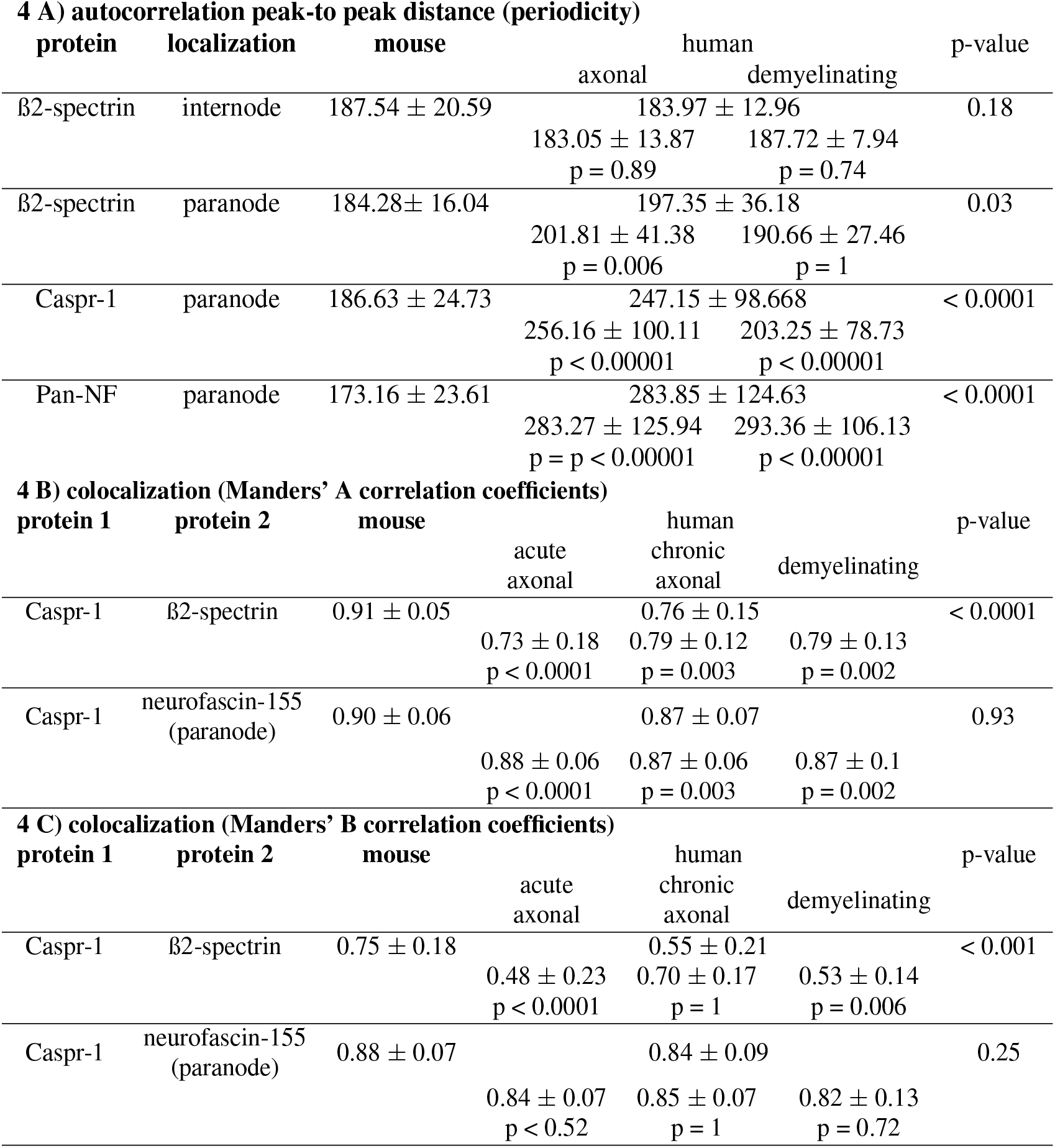
*d*STORM imaging parameters in mice and patients with polyneuropathies. (A) Autocorrelation peak-to peak distances (periodicity) of internodal and paranodal ß2-spectrin, Caspr-1 and neurofascin-155 in mice and patients with polyneuropathies. (B-C): Colocalization analysis using Manders’ A and B correlation coefficients between Caspr-1 with ß2-spectrin and with the paranodal fraction of pan-neurofascin (neurofascin-155) are displayed in mouse and human Nodes of Ranvier. P-values of Kruskal-Wallis test with Dunn’s correction for multiple testing are shown in the right column (mouse vs. human) and below the individual results of subanalysis (mouse vs. human axonal / demyelinating neuropathy subcohort).

**Table S5.** Morpometics at the Node: High content confocal imaging and analysis data in single patients. Means ± standard deviations and medians of nodo-paranodal parameters (nodes / tissue volume, axonal paranodal diameter, nodo-paranodal length, nodal length) are shown for single patients of the cohort. Adjusted p-values are displayed compared to intact human nerves using Kruskal-Wallis test with Dunn’s correction for multiple testing. Patient biopsies that were used to select intact nodes are marked with asterisks. Abbreviations: no = number, SD standard deviation.

| no | n of measured nodes | nodal count | axonal paranodal diameter |  |  | nodo-paranodal length |  |  | nodal length |  |  |
| --- | --- | --- | --- | --- | --- | --- | --- | --- | --- | --- | --- |
| | | | mean ± SD ($\mu\text{m}$) | median ($\mu\text{m}$) | adjusted p-value | mean ± SD ($\mu\text{m}$) | median ($\mu\text{m}$) | adjusted p-value | mean ± SD ($\mu\text{m}$) | median ($\mu\text{m}$) | adjusted p-value |
| 1 | 40 | 16.91 | 2.74 ± 1.12 | 2.57 | 0.1072 | 13.09 ± 5.51 | 11.45 | 0.0006 | 1.17 ± 1.58 | 0.89 | >0.9999 |
| 2* | 40 | 21.53 | 2.85 ± 0.75 | 2.85 | 0.0004 | 14.11 ± 6.45 | 12.30 | <0.0001 | 0.99 ± 0.81 | 0.79 | >0.9999 |
| 3 | 40 | 18.32 | 2.16 ± 0.65 | 2.23 | >0.9999 | 13.88 ± 7.74 | 10.40 | 0.0016 | 1.16 ± 0.33 | 1.10 | 0.1692 |
| 4 | 26 | 12.17 | 2.40 ± 0.96 | 2.46 | >0.9999 | 12.21 ± 2.91 | 11.26 | 0.0032 | 0.91 ± 0.60 | 0.84 | >0.9999 |
| 5 | 39 | 14.17 | 1.80 ± 0.66 | 1.64 | 0.1171 | 14.36 ± 6.66 | 12.25 | <0.0001 | 1.23 ± 0.81 | 1.06 | 0.5359 |
| 6 | 37 | 13.22 | 2.08 ± 0.73 | 1.93 | >0.9999 | 24.66 ± 17.64 | 15.99 | <0.0001 | 2.16 ± 2.49 | 1.45 | <0.0001 |
| 7 | 40 | 38.96 | 2.41 ± 0.95 | 2.26 | >0.9999 | 12.99 ± 9.33 | 9.88 | 0.3935 | 1.73 ± 3.96 | 0.86 | >0.9999 |
| 8* | 40 | 30.12 | 2.16 ± 0.84 | 1.94 | >0.9999 | 12.11 ± 6.11 | 10.65 | 0.0487 | 0.81 ± 0.49 | 0.70 | >0.9999 |
| 9 | 22 | 18.28 | 2.61 ± 1.01 | 2.32 | >0.9999 | 16.43 ± 10.16 | 13.16 | 0.0002 | 1.07 ± 0.83 | 0.78 | >0.9999 |
| 10 | 38 | 28.67 | 2.26 ± 0.93 | 2.01 | >0.9999 | 10.90 ± 3.35 | 10.74 | 0.2976 | 0.85 ± 0.34 | 0.78 | >0.9999 |
| 11 | 40 | 36.37 | 2.03 ± 0.65 | 1.95 | >0.9999 | 13.65 ± 8.22 | 11.53 | 0.0012 | 0.92 ± 1.59 | 0.59 | 0.2211 |
| 12* | 40 | 47.41 | 1.79 ± 0.72 | 1.48 | 0.0652 | 11.29 ± 3.23 | 10.56 | 0.1324 | 0.92 ± 0.74 | 1.48 | <0.0001 |
| 13 | 35 | 22.31 | 2.39 ± 0.93 | 2.23 | >0.9999 | 14.96 ± 2.39 | 11.79 | <0.0001 | 0.93 ± 0.59 | 0.76 | >0.9999 |
| 14* | 40 | 19.95 | 2.74 ± 1.14 | 2.57 | 0.334 | 10.46 ± 7.81 | 9.47 | >0.9999 | 0.93 ± 0.93 | 0.71 | >0.9999 |
| 15 | 40 | 21.80 | 3.14 ± 1.11 | 3.21 | 0.0001 | 11.49 ± 3.08 | 10.73 | 0.0231 | 1.32 ± 0.40 | 1.27 | 0.0008 |
| 16 | 30 | 82.68 | 2.86 ± 1.12 | 2.89 | 0.0651 | 14.00 ± 6.67 | 11.53 | 0.0008 | 1.16 ± 1.75 | 0.65 | >0.9999 |
| 17 | 235 | 32.51 | 2.24 ± 0.58 | 2.20 | >0.9999 | 14.35 ± 8.26 | 12.80 | <0.0001 | 1.23 ± 0.52 | 1.16 | 0.0727 |

**Table S6.**
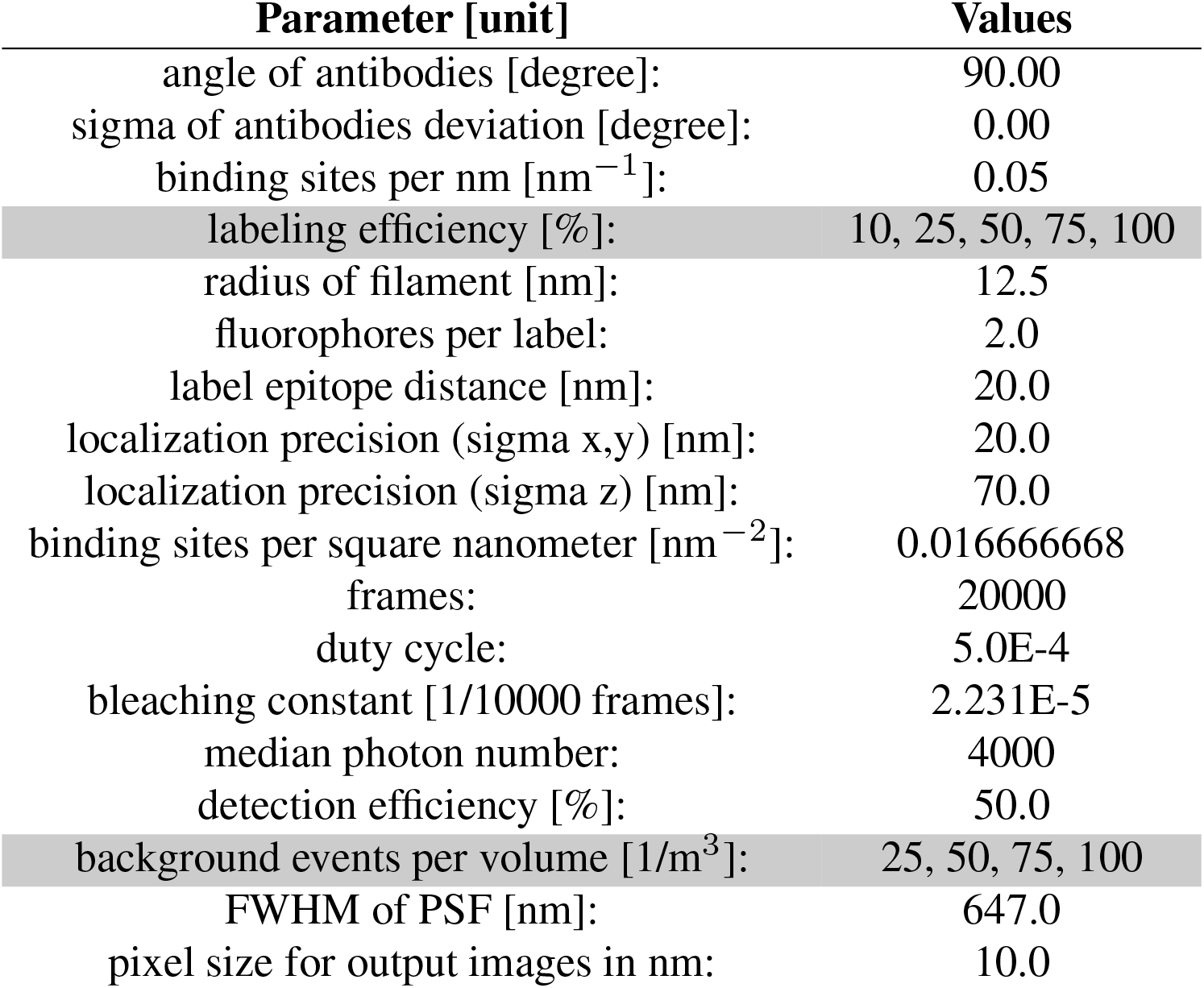
Summary of the parameters for the simulation of SMLM datasets of ground truth periodic line patterns with SuReSim. The parameter sets used in the sweep are highlighted in gray.

| Parameter [unit] | Values |
| --- | --- |
| angle of antibodies [degree]: | 90.00 |
| sigma of antibodies deviation [degree]: | 0.00 |
| binding sites per nm [$\text{nm}^{-1}$]: | 0.05 |
| labeling efficiency [%]: | 10, 25, 50, 75, 100 |
| radius of filament [nm]: | 12.5 |
| fluorophores per label: | 2.0 |
| label epitope distance [nm]: | 20.0 |
| localization precision (sigma x,y) [nm]: | 20.0 |
| localization precision (sigma z) [nm]: | 70.0 |
| binding sites per square nanometer [$\text{nm}^{-2}$]: | 0.016666668 |
| frames: | 20000 |
| duty cycle: | 5.0E-4 |
| bleaching constant [1/10000 frames]: | 2.231E-5 |
| median photon number: | 4000 |
| detection efficiency [%]: | 50.0 |
| background events per volume [$1/\text{m}^3$]: | 25, 50, 75, 100 |
| FWHM of PSF [nm]: | 647.0 |
| pixel size for output images in nm: | 10.0 |

**Table S7.** Antibodies used for fluorescence stainings

| target protein<br>(conjugate) | host | dilution | company | catalogue number |
| --- | --- | --- | --- | --- |
| pan-neurofascin | polyclonal chicken IgY | 0.4 $\mu\text{g}/\text{ml}$ | R & D Systems | AF3235 |
| Caspr-1 | polyclonal rabbit IgG | 0.2 $\mu\text{g}/\text{ml}$ | Abcam | ab34151 |
| b2-Spectrin II | monoclonal mouse IgG | 5 $\mu\text{g}/\text{ml}$ | BD Biosciences | 612563 |
| Myelin basic protein | monoclonal rat IgG | 10 $\mu\text{g}/\text{ml}$ | Abcam | ab7349 |
| chicken IgY(Alexa 647) | polyclonal goat IgG | 10 $\mu\text{g}/\text{ml}$ | Invitrogen | A-21449 |
| rabbit IgG (Alexa532) | polyclonal goat IgG | 10 $\mu\text{g}/\text{ml}$ | Invitrogen | A-11009 |
| rat IgG (Alexa488) | polyclonal donkey IgG | 3.75 $\mu\text{g}/\text{ml}$ | Jackson IR | 712-545-150 |
| rabbit IgG (Cy3) | polyclonal donkey IgG | 5 $\mu\text{g}/\text{ml}$ | Jackson IR | 711-165-152 |

**Table S8.** Customized two-color *d*STORM microscope specifications.

|  |  |
| --- | --- |
| **Illumination** | 640 nm laser (iBEAM-smart-640-S, Toptica)<br>532 nm laser (GEM 532 nm, 250 mW, Laser Quantum) |
| **Clean up filters** | 640/8 nm MaxDiode™ laser clean-up, Semrock<br>Filter ZET 532/10 clean-up, Chroma |
| **Dichroic filter** | QuadLine Laser-Beamsplitter R405/488/532/635, Semrock<br>Quad-Notch Filter 405/488/532/635, Semrock |
| **Two-channel detection** | Optosplit II (Crain) with Beamsplitter T 635 LPXR, Semrock |
| **Emission filters** | BrightLine HC 593/40, Semrock<br>700/75 ET, Chroma |
| **Camera** | iXon ultra DU-897U-CSO-BV, Andor |

**Table S9.**
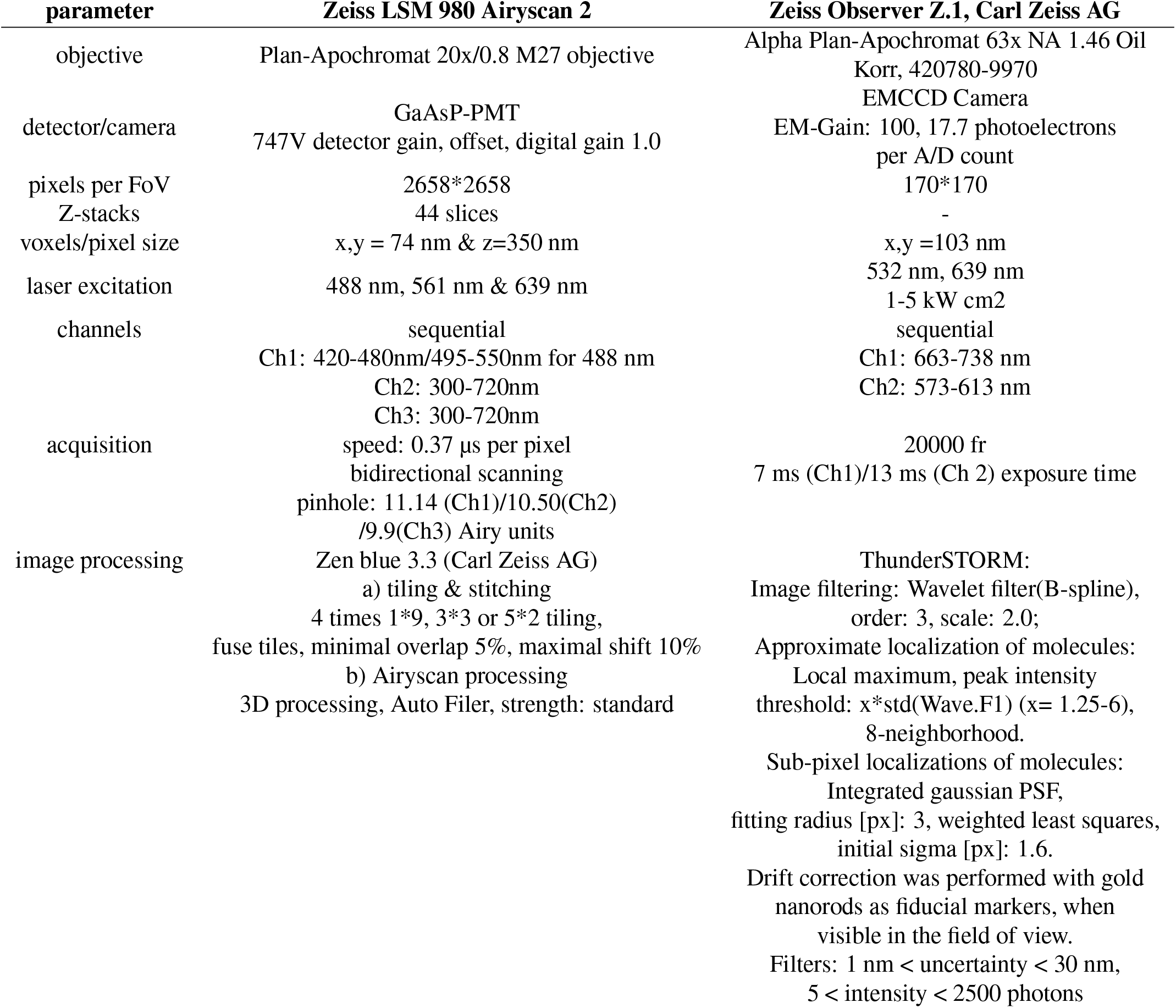
Acquisition parameters and image processing details. Abbreviations: n/a: not applicable.

| **parameter** | **Zeiss LSM 980 Airyscan 2** | **Zeiss Observer Z.1, Carl Zeiss AG** |
| --- | --- | --- |
| objective | Plan-Apochromat 20x/0.8 M27 objective | Alpha Plan-Apochromat 63x NA 1.46 Oil Korr, 420780-9970 |
| detector/camera | GaAsP-PMT<br>747V detector gain, offset, digital gain 1.0 | EMCCD Camera<br>EM-Gain: 100, 17.7 photoelectrons per A/D count |
| pixels per FoV | 2658*2658 | 170*170 |
| Z-stacks | 44 slices | - |
| voxels/pixel size | x,y = 74 nm & z=350 nm | x,y =103 nm |
| laser excitation | 488 nm, 561 nm & 639 nm | 532 nm, 639 nm<br>1-5 kW cm<sup>2</sup> |
| channels | sequential<br>Ch1: 420-480nm/495-550nm for 488 nm<br>Ch2: 300-720nm<br>Ch3: 300-720nm | sequential<br>Ch1: 663-738 nm<br>Ch2: 573-613 nm |
| acquisition | speed: 0.37 μs per pixel<br>bidirectional scanning<br>pinhole: 11.14 (Ch1)/10.50(Ch2)<br>/9.9(Ch3) Airy units | 20000 fr<br>7 ms (Ch1)/13 ms (Ch 2) exposure time |
| image processing | Zen blue 3.3 (Carl Zeiss AG)<br>a) tiling & stitching<br>4 times 1*9, 3*3 or 5*2 tiling,<br>fuse tiles, minimal overlap 5%, maximal shift 10%<br>b) Airyscan processing<br>3D processing, Auto Filer, strength: standard | ThunderSTORM:<br>Image filtering: Wavelet filter(B-spline),<br>order: 3, scale: 2.0;<br>Approximate localization of molecules:<br>Local maximum, peak intensity<br>threshold: $x \cdot \text{std}(\text{Wave.F1})$ ($x = 1.25-6$),<br>8-neighborhood.<br>Sub-pixel localizations of molecules:<br>Integrated gaussian PSF,<br>fitting radius [px]: 3, weighted least squares,<br>initial sigma [px]: 1.6.<br>Drift correction was performed with gold nanorods as fiducial markers, when visible in the field of view.<br>Filters: 1 nm < uncertainty < 30 nm,<br>5 < intensity < 2500 photons |

## Supplementary Figure 1-4

**Fig. S1.**
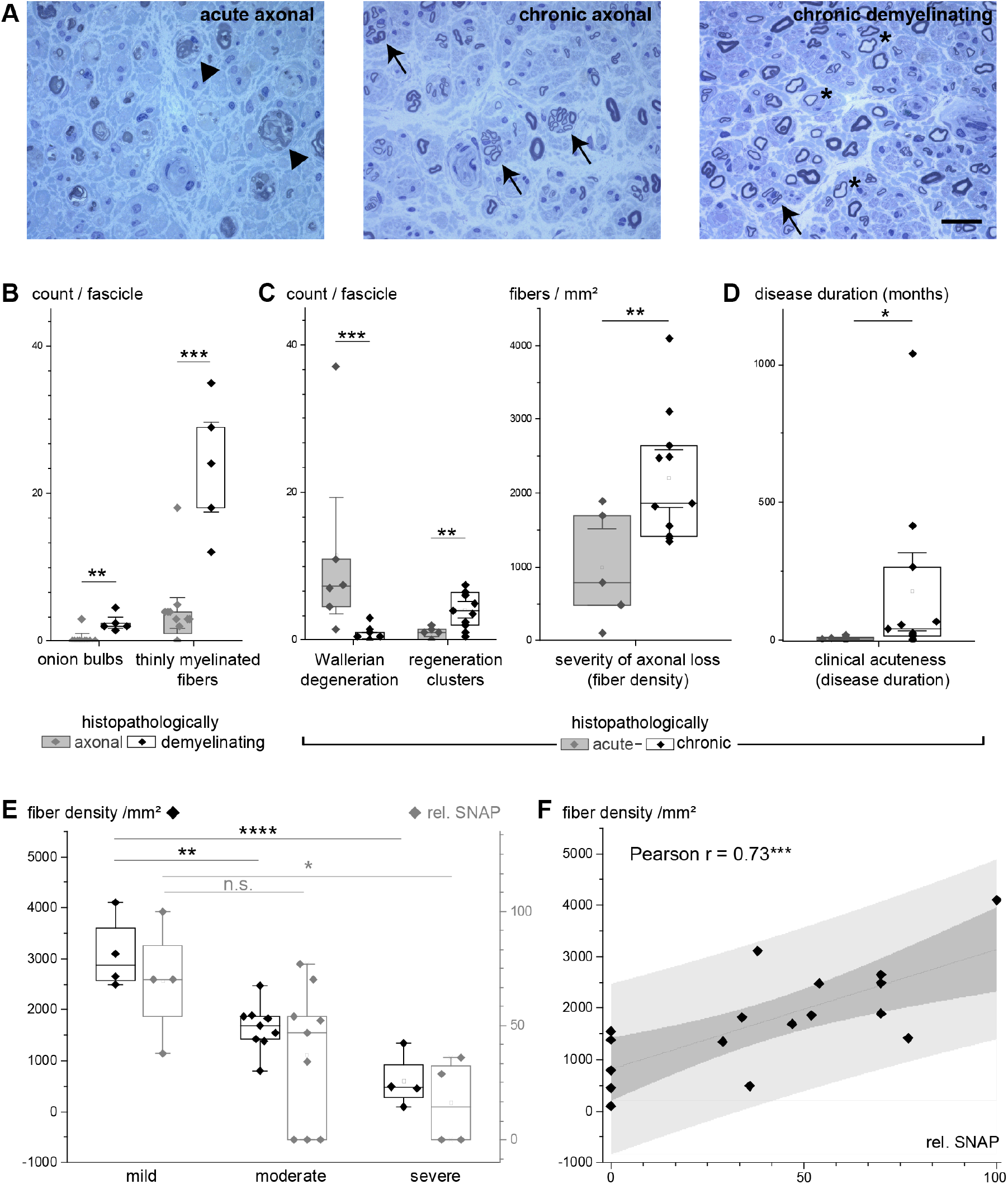
Histopathological assessment of cross-sectional sensory nerve biopsies and validation. (A) Sensory nerve semithin cross sections with toluidine blue staining in acute axonal neuropathy, chronic axonal neuropathy and demyelinating neuropathy (left to right) show axonal loss with actively degeneration fibers (arrowheads, left) in acute axonal neuropathy, clusters of regenerating fibers in chronic axonal and demyelinating neuropathy (arrows) and thinly myelinated fibers in demyelinating neuropathy (asterisks). Scale bar = 20μm. (B) Histopathological classification by type of disease was validated by comparing the counts per fascicle (y-axis) of onion bulbs and thinly myelinated fibers (x-axis) between biopsies rated as axonal (gray) and demyelinating (black/white, see legend) using Mann-Whitney test. (C) Histopathological classification by acuteness was validated by comparing the counts / fascicle (left graph, y-axis) of Wallerian degeneration figures and regeneration clusters (left graph, x-axis) between biopsies rated as acute (gray) and chronic (black/white, see legend). Fiber density /mm^2^ as an indicator of severity of axonal loss (right graph, y-axis) was significantly reduced in acute vs. chronic neuropathy (Student’s t test). (D) Histopathological classification was clinically validated by comparing the clinical disease duration in acute vs. chronic neuropathy using Mann-Whitney test. (E-F) Histopathologic classification by severity was validated by nerve conduction studies data (exact values see Supplemental Table 3). (E) Fiber density /mm^2^ (left y-axis, black) and relative SNAP in percent of the age-adjusted lower limit of normal (right y-axis, gray) significantly differ according to the severity of axonal loss, assessed by one-way ANOVA with Bonferroni adjustment (fiber density) and Welch one way ANOVA with Dunnett’s T3 multiple comparison test (SNAP). (F) Relative SNAP (x-axis) correlated positively to the fiber density (y-axis) using Pearson’s correlation coefficient. Significance level: * p < 0.05. ** p < 0.01, *** p < 0.001, **** p < 0.0001. Abbreviations: n.s. = not significant, SNAP = sensory nerve action potential.

**Fig. S2.**
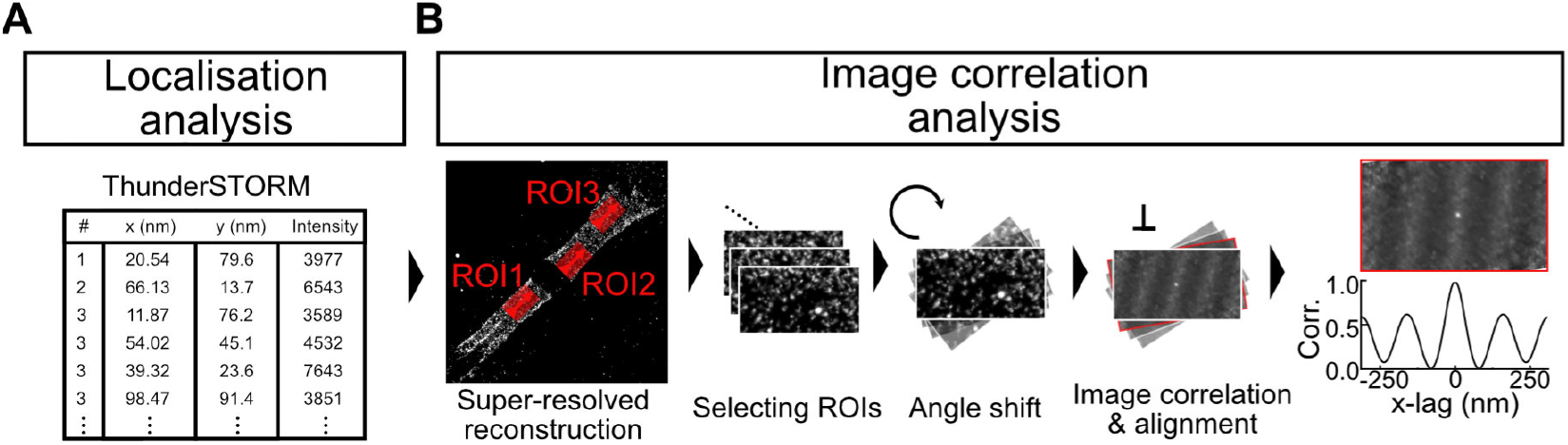
Image reconstruction and analysis workflow. (A) The ThunderSTORM localisation analysis of the SMLM image sequences provides a list of molecule positions. Based on this list, a super-resolved image of ultrastructural organization of the protein of interest at the node of Ranvier is reconstructed. (B) The Image correlation analysis is performed with a custom MatLab tool which allows to select regions of interest in the super-resolved image. The ROIs are rotated in steps before image auto-correlation to align the pattern perpendicular to the horizontal axis.

**Fig. S3.**
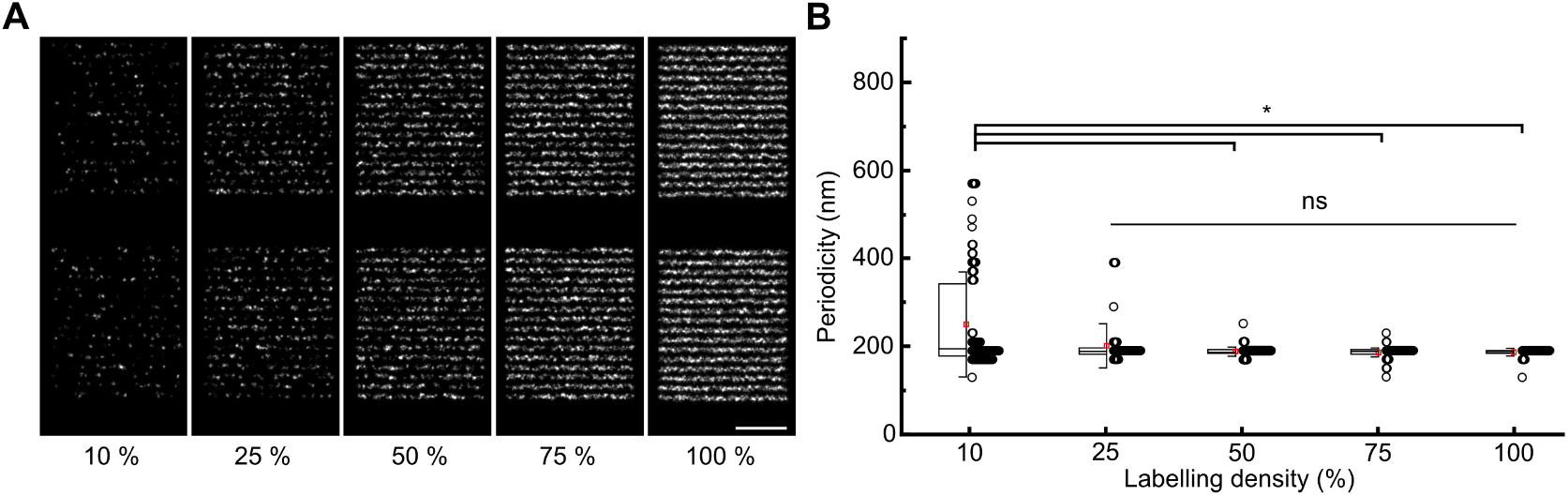
Super-resolution image autocorrelation analysis robustly identifies structural periodicity in the case of incomplete labeling. (A) Based on super-resolved reconstructions of simulated SMLM data of a ground truth line structure with 190 nm periodicity and labeling densities from 10 to 100%, autocorrelation (B) successfully retrieves the periodicity spacing at minimum 25% (median ± standard deviation; 10%:194 ± 119 nm, n = 91; 25%: 188 ± 50 nm, n = 94; 50%: 186 ± 11 nm, n = 131; 75%: 188 ± 11 nm, n = 150; 100%: 188 ± 9 nm, n = 134; Kruskal-Wallis test). Scale bar 1 μm.

**Fig. S4.**
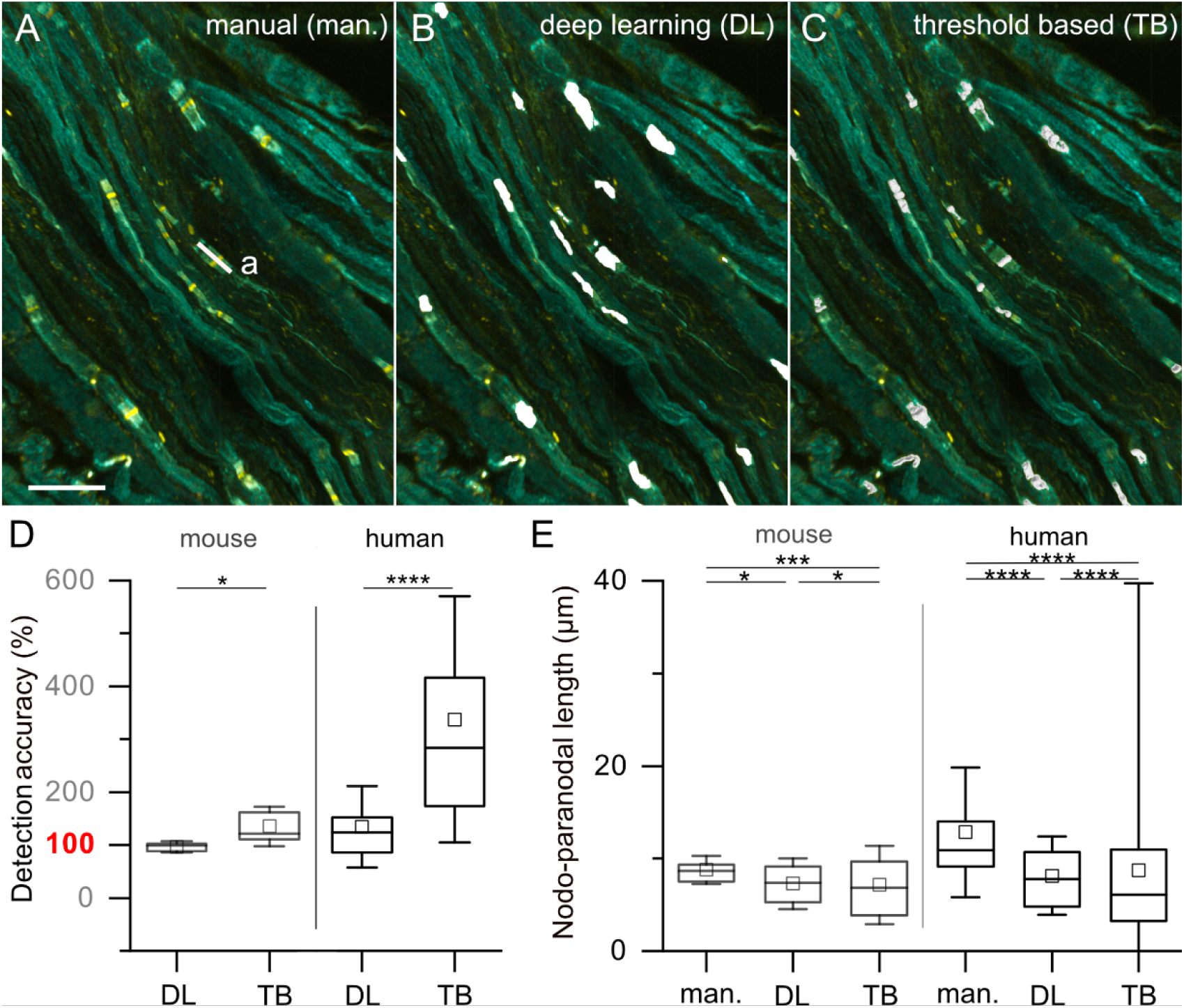
Assessment of node detection and nodo-paranodal length by deep learning based segmentation vs. threshold based segmentation. (A-C) Murine ischiadic nerve teased fiber preparations with two-color labeling (pan-neurofascin in yellow and Caspr-1 in cyan, A) were segmented using deep learning (DL, B) or threshold based segmentation using Imaris Software (TB, C) to identify single nodes of Ranvier. Scale bar = 20 μm. (D) Detection and counting accuracy of the nodes of Ranvier using DL vs TB. In both mouse and human samples DL performed significantly better than TB in detecting nodes of Ranvier, using manual assessment as a reference. Median ± std deviation in mouse was 0.99 ± 0.1 vs 1.21 ± 1.36, p = 0.046 and in human 1.21 ± 0.77 vs 2.83 ± 2.33 p < 0.0001, determined by Kruskal-Wallis ANOVA with adjustment for multiple testing. (E) Nodo-paranodal length calculated from DL and Imaris threshold based image segmentation was compared to manual ROI definition using FIJI by two independent and blinded investigators. Data were compared in murine and human samples. In both, statistically significant differences between automated and manual analysis could be detected. In both cases, results from image segmentation with DL were closer to manual assessment than threshold based assessment (median ± std deviation nodo-paranodal length in μm in mouse: man.: 8.6 ± 1.5, DL: 7.2 ± 2.7 p = 0.02, TB: 6.9 ± 4.2 p < 0.0001, human: man.: 10.9 ± 7.0, DL: 7.8 ± 4.2 p < 0.001, TB: 6.1 ± 31.0 p < 0.0001, calculated by Kruskal-Wallis ANOVA with adjustment for multiple testing. Significance level: * p < 0.05. ** p < 0.01, *** p < 0.001, **** p < 0.0001.

## Supplementary Movies

**Supplementary Movie M1: Assessment of nodo-paranodal parameters based on multicolor large field of view fluorescence imaging**. Human sural nerve teased fiber preparations with triple-labeling (MBP in magenta, pan-neurofascin in yellow and Caspr-1 in cyan) were imaged to asses the nodo-paranodal length, paranodal diameter and nodeal gap as indicated in the video. Scale bar = 100 μm.

